# Highly Accurate Non-Invasive Preimplantation Genetic Testing for Monogenic and Polygenic Diseases from Spent Medium

**DOI:** 10.1101/2024.11.10.24317049

**Authors:** Lei Huang, Jin Huang, Minyue Ma, Yangyun Zou, Ruiqi Zhang, Guangjun Yin, Quangui Wang, Yingying Xia, Jialin Jia, Zeyu Wu, Dandan Cao, Weiliang Song, Yaqiong Tang, Kai Liu, Xiaoran Chai, Guo-Bo Chen, Sijia Lu, Hongmei Peng, Hao Ge, Jie Qiao, X. Sunney Xie

## Abstract

Traditional preimplantation genetic testing for in vitro fertilization (IVF) requires invasive trophectoderm biopsy, which may harm the embryo. Non-invasive preimplantation genetic testing (niPGT) utilizing cell-free DNA from spent embryo culture medium (SCM) has been attempted, but trace DNA content, maternal cell contamination, and high allele drop-out rates are barriers. Here we report an niPGT methodology achieving high diagnostic accuracy among reportable samples across diverse embryo and disease contexts. Reengineered linear whole-genome amplification with an extremely low error rate is followed by application of a Bayesian algorithm designed to determine which parental haplotypes are inherited at disease-causing sites. Among 191 SCM samples from 29 families affected by rare monogenic disorders, our method amplified all samples and enabled diagnosis of 220 out of 277 pathogenic alleles. All diagnoses (220/220) were concordant with invasive biopsy control. For select embryos, personalized genome-wide genotyping enabled exploratory polygenic risk assessment for Type II Diabetes. Our methodology requires no modification to IVF workflows and represents a key technological advance toward scalable, accurate, and safe niPGT for both monogenic and polygenic diseases.

## Introduction

Preimplantation genetic testing (PGT) is widely used in clinical in vitro fertilization [1] to screen embryos for monogenic diseases [2,3], aneuploidy [4,5], or structural rearrangements [6,7]. PGT based on next-generation sequencing allows simultaneous analysis of chromosome copy number, linkage to causal or risk alleles, and targeted haplotyping for specific mutations [8]. However, current PGT protocols require trophectoderm biopsy, which may pose risks to embryo health and is prone to sampling bias due to embryo mosaicism [9–11]. Recently, minimally invasive or non-invasive methods using cell-free DNA from blastocoele fluid [12] or spent embryo culture medium (SCM) [13–15] have been reported as lower-risk alternatives. Despite their potential, major challenges remain. Trace DNA and a wide variation in DNA fragment length have led to highly variable success rates in published studies from different clinical centers [15–18]. Further, DNA amplified from SCM may be contaminated by cell-free maternal DNA fragments [19], which are often longer than embryonic DNA fragments [20,21] and are therefore preferentially amplified. The presence of high levels of human serum albumin protein in culture medium is also known to cause amplification difficulties.

Current commercial single-cell whole-genome amplification kits use amplification methods that have not been specially optimized to address these challenges. Commonly used methods include Degenerate Oligonucleotide-Primed PCR (DOP-PCR), Multiple Displacement Amplification (MDA), or Multiple Annealing and Looping Based Amplification Cycles (MALBAC). In 2017, Linear Amplification via Transposon Insertion (LIANTI) was developed [22], significantly reducing the false positive rate of single nucleotide variation detection, with much less fragment-length amplification bias. However, the published LIANTI amplification protocol often fails to amplify DNA from SCM.

An additional challenge for niPGT is performing linkage analysis to track the inheritance of parental haplotypes carrying monogenic disease-causing variants. Linkage analysis has become a standard approach to circumvent false-positive and false-negative single-nucleotide variants (SNVs) that can arise during whole-genome amplification. Traditional linkage analyses typically require at least two fully informative markers near the mutation site, and assume allele drop-out rates below 5% [15,18,23]. However, allele drop-out rates in SCM or blastocoele fluid often exceed 5%, meaning reliance on a few informative sites for linkage analysis may lead to misdiagnosis, while at the same time reliance on SNP markers far from the mutation site increases recombination risk. Maternal cell contamination (MCC) can also raise the misdiagnosis rate. To date, these obstacles have prevented traditional linkage analysis methods from being successfully applied to niPGT.

Given these challenges, it is perhaps not surprising that while there are many reports of non-invasive PGT (niPGT) for aneuploidy [13,14,24,25], application to monogenic disorders (niPGT-M) [15,16,17,26–29] has yielded mixed results, with genotype concordance to trophectoderm biopsy varying from 21% to 88% [15–18]. Clinically, diagnostic accuracy is the foremost priority in PGT for monogenic disorders. While niPGT success rates are generally reported to be higher using SCM as opposed to blastocoele fluid [16,29], neither cell-free source is close to achieving the 99% concordance with embryonic cell results that is typically required for PGT-M clinical diagnostics [30].

PGT also holds promise for assessing the risk of polygenic disorders such as coronary heart disease, diabetes, and cancers [31–33]. A polygenic risk score (PRS) quantifies the cumulative effects of many common variants and therefore represents probabilistic risk estimates rather than deterministic diagnoses. Across multiple diseases, studies have shown that hypothetical selection of a sibling with a lower PRS would result in significant relative-risk reductions [31]. While fully acknowledging the ethical and societal questions surrounding PGT-P, non-invasive PGT for polygenic disorders (niPGT-P) could help to facilitate scientific evaluation of potential benefits while reducing clinical risks [34]. However, the limited amount and poor quality of cell-free DNA have prevented the use of niPGT-P for PRS evaluation.

The aim of this study was to improve DNA amplification efficiency and diagnostic accuracy in non-invasive PGT for monogenic and polygenic disorders to a level approaching clinical applicability. We reengineered the LIANTI method to amplify trace DNA from SCM, overcoming challenges posed by high protein concentration in samples and amplification biases arising from DNA fragment-size differences. We introduced a Bayesian linkage analysis method (BASE-niPGT-M) that explicitly models high allele drop-out and maternal cell contamination (MCC), which accurately detects disease-carrying alleles and provides a confidence level for each monogenic diagnosis. We also employed a Pedigree-Population-based Imputation with Haploid Assumption (PPIHA) method to reconstruct the embryonic genome from fragmented raw SCM genome data. The resulting high coverage and accuracy of the genome-wide genotyping enabled calculation of polygenic risk scores for Type II Diabetes. Together, these advances overcome the major technical barriers that have long hindered the accuracy of niPGT for monogenic and polygenic disorders, establishing a reliable and accurate protocol that lays the foundation for clinical translation.

## RESULTS

### Characterization of Cell-Free DNA in Spent Embryo Culture Medium by a Reengineered LIANTI Method

Unlike biopsy samples, which typically contain five to eight complete cell nuclei, SCM contains very low amounts of DNA (≤20 pg, commonly ≤6 pg)[13,19]. Further, our SCM samples exhibited DNA fragments of approximately 180-200 base pairs and their multiples, a hallmark of internucleosomal cleavage [35,36], posing a significant challenge for DNA amplification (Extended Data Fig. 1 and Supplementary Methods). Using DNA from cell lines, we compared the fragment-size amplification biases of the current genome amplification methods for PGT and found that LIANTI exhibited the least bias (short:long = 1:5), while MDA and MALBAC showed a stronger preference for amplifying longer fragments, with short:long amplification biases of 1:500 and 1:6000, respectively (Table S1 and Supplementary Methods).

However, the original LIANTI method was rarely successful in amplifying the minute quantities of DNA in SCM. We reengineered the LIANTI method by modifying the lysis step, adding more transposons for insertion, introducing DNA primers during first-strand DNA synthesis, and implementing exponential amplification during second-strand synthesis (Extended Data Fig. 2 and Supplementary Methods). The reengineered LIANTI method achieved a 100% amplification success rate for all the clinical samples and demonstrated high SNP detection accuracy (Extended Data Fig. 3 and Table S2). The average direct genome coverage for cell-free DNA from SCM collected from Clinical Center A was 35.0% (Median 33.5%, STDev 17.8%, Interquartile range [IQR] 23.6%-46.7%) (Extended Data Fig. 4), in contrast to the genome coverage of biopsy samples, which typically exceeds 95% [30]. This lack of coverage is not primarily due to amplification-induced allele drop-out, which is characterized by short tracts of missing data, but rather to missing template DNA in the medium itself, which is characterized by large contiguous blocks of missing information (Extended Data Fig. 5). This phenomenon often results in the loss of informative SNPs used in traditional linkage analysis, both upstream and downstream of the pathogenic site.

To estimate the fraction of maternal DNA contamination (synonymous here with MCC level), we used SNPs that were homozygous but different in each parental genotype (for example A/A in one parent, and B/B in the other), specifically within a 20 Mb region centered on each pathogenic locus. Using a scale ranging from –100% (all paternal) to +100% (all maternal) where a value of zero represents balanced maternal and paternal allele contributions, we observed values ranging from -55.1% to 99.7% for samples from Clinical Center A (Extended Data Fig. 6). Employing the iterative estimation from our Bayesian linkage analysis method (see below and Methods), we were able to correct for maternal DNA contamination levels ranging from 1% to 99.7% (Supplementary Methods, Table S3, Extended Data Fig. 6). Among all the 182 autosomal disease samples from both Clinical Centers A and B, MCC levels were minimal (<2%) for 88 samples, low (2%–30%) for 66, moderate (30%-80%) for 18, and high (>80%) for 10 (Table S3).

### Non-invasive PGT for Monogenic Disorders

As part of the niPGT-M workflow (Fig. 1 and Methods), data collection was divided into two stages (Fig. 1A). In the first stage (pre-phasing), sequencing was performed using DNA from the father and mother of the embryo and from any living siblings of the embryo, and alternatively or in addition, from parental siblings, grandparents, or discarded embryos. The sequencing data were used to reconstruct, for each parent, the haplotypes inherited from their own mother and father. In the second stage (genetic information acquisition), the DNA in SCM was collected and amplified using a reengineered LIANTI method, followed by sequencing and SNP calling.

**Figure 1.**
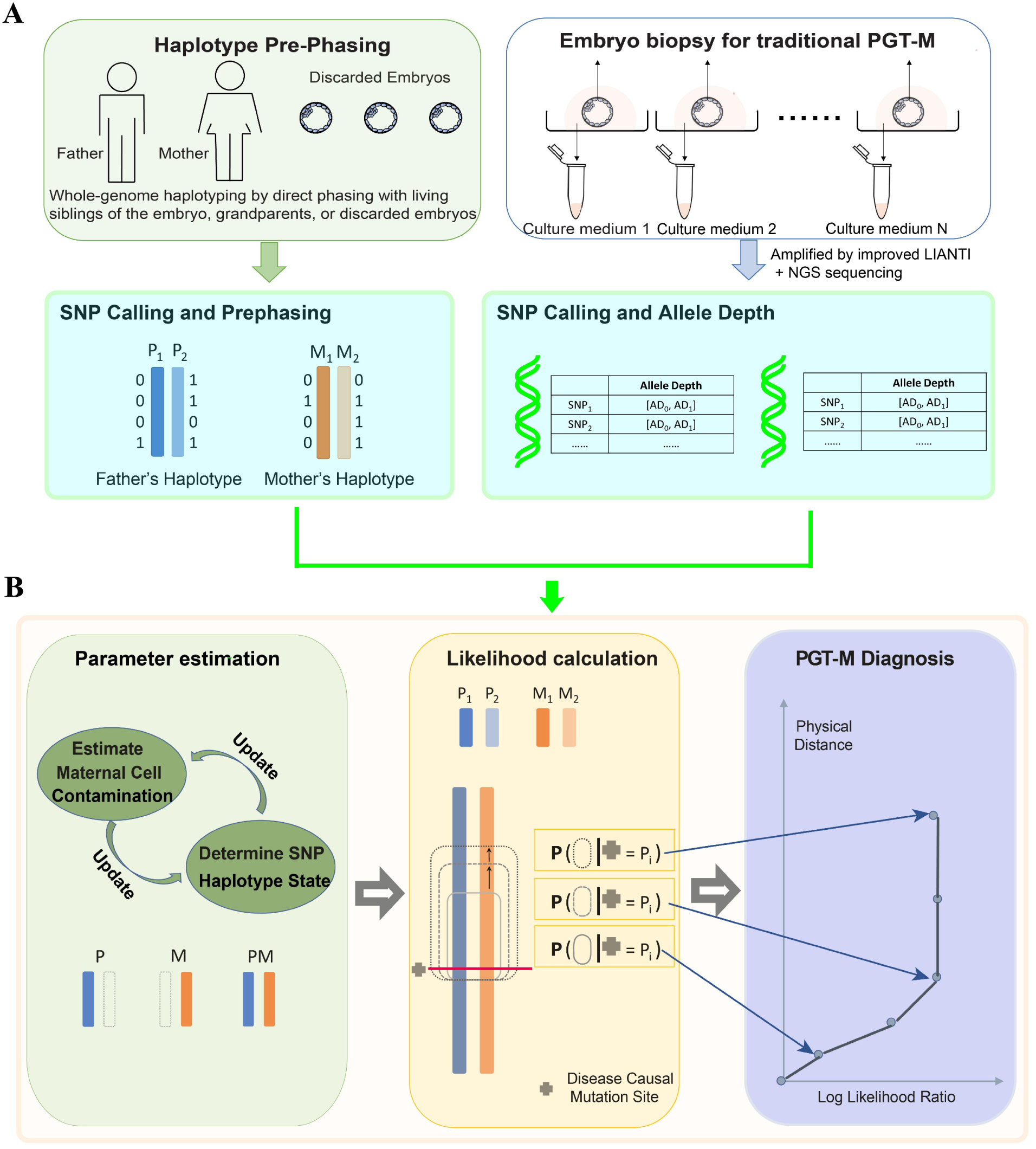
niPGT-M workflow using spent embryo culture medium. **A.** (Left) In pre-phasing, sequencing was performed using DNA from the father and mother of the embryo and from living siblings of the embryo, and alternatively or in addition, from parental siblings, grandparents, or discarded embryos. (Right) Fertilized embryos were cultured, and immediately after transferring the blastocysts to another dish for biopsy, culture media were removed for analysis. For comparison, trophectoderm cells from each blastocyst were biopsied and underwent traditional PGT for monogenic disorders. All SCM samples underwent reengineered LIANTI protocol genome amplification and next-generation sequencing, followed by SNP calling. Allele depth refers to the number of reads mapped to a specific SNP. **B.** SNP data from SCM samples, together with the phased haplotypes of the parents, are input to the Bayesian linkage analysis method. After iteratively estimating the levels of maternal cell contamination and haplotype status of each SNP, the log-likelihood ratio of inheriting the pathogenic allele versus the non-pathogenic allele was calculated. The diagnosis for monogenic disorders is based on the log-likelihood ratio curve as a function of distance to the disease-causing mutation site.

For analysis of these data, we developed BASE-niPGT-M (BAyesian linkage analySis mEthod for non-invasive Preimplantation Genetic Testing of Monogenic disorders, Fig. 1B, Methods, Extended data Fig. 7 and Supplementary Methods). This computational approach addresses the challenges posed by maternal cell contamination and low coverage in SCM samples. The method builds upon a Hidden Markov Model (HMM) structure, as employed in several previous linkage-analysis approaches designed for high-quality genotyping data with low error and dropout rates [37,38]. Our model explicitly incorporates the empirically estimated allele drop-out and maternal contamination rates, allowing it to remain robust under the highly sparse and error-prone cell-free DNA (cfDNA) data conditions. Unlike previous approaches for high-quality sequencing data, which often use a predetermined number of SNPs for linkage analysis, our model incrementally incorporates SNPs, starting from the disease-causing mutation site. This process allows us to calculate the log-likelihood ratio of inheriting disease-carrying chromosomes versus disease-free chromosomes for each SNP subset, ultimately resulting in a log-likelihood ratio curve (Fig. 1B, Extended data Fig. 7 and Supplementary Methods). The distinct characteristics of this curve enabled us to classify the results into four confidence categories: High (posterior probability ≥99.9%), Moderate (posterior probability ≥95%), Low (diagnosis is supported but with a posterior probability < 95%), and Undetermined (see Methods).

We selected one case (Family 2) to illustrate the niPGT-M diagnose pathway described here, from the derivation of SNP distribution from each cell-free DNA sample to the final diagnoses. This case involved an autosomal dominant genetic disease, where the father carried a genetic variant for Marfan syndrome, a heritable connective tissue disorder. This disorder is known for its characteristic features such as elongated limbs, aortic aneurysms, and mitral valve prolapse. Genetic diagnosis of the father revealed a mutation c.643C>T at the FBN1 gene, known to cause Marfan syndrome. Eight embryos from the couple developed to the blastocyst stage, and several trophectoderm cells were biopsied from each embryo for independent traditional PGT. All eight SCM samples were collected, and cell-free DNA was amplified using the reengineered LIANTI method. The linkage analysis results of these eight samples are shown in Fig. 2A. For samples #1 and #2, the detected SNP distributions were sparse, with long intervals between informative SNPs (Fig. 2A). Further, the SNPs flanking the causal mutation site indicated different haplotypes from the father, implying a recombination event near the causative allele, leading to the classification of samples #1 and #2 as ’Undetermined’ (Fig. 2B). In contrast, samples #3-8 showed dense SNP distributions, leading to a determination of 4 disease-carrying and 2 disease-free haplotypes. Among these 6 samples (#3-8), all SNPs upstream and downstream of the causal mutation site indicated the same haplotype (Fig. 2B) resulting in a ’High confidence’ classification, with the exception of sample #7. In sample #7, the detected SNPs were dense and the inferred haplotypes upstream and downstream of the causal mutation site were concordant, except for a single SNP located relatively close to the causal site. This led to a ’Moderate confidence’ label due to our conservative diagnosis classification rules (Fig. 2B, Supplementary Methods). Each of the six diagnoses was consistent with the diagnosis made using trophectoderm biopsies.

**Figure 2.**
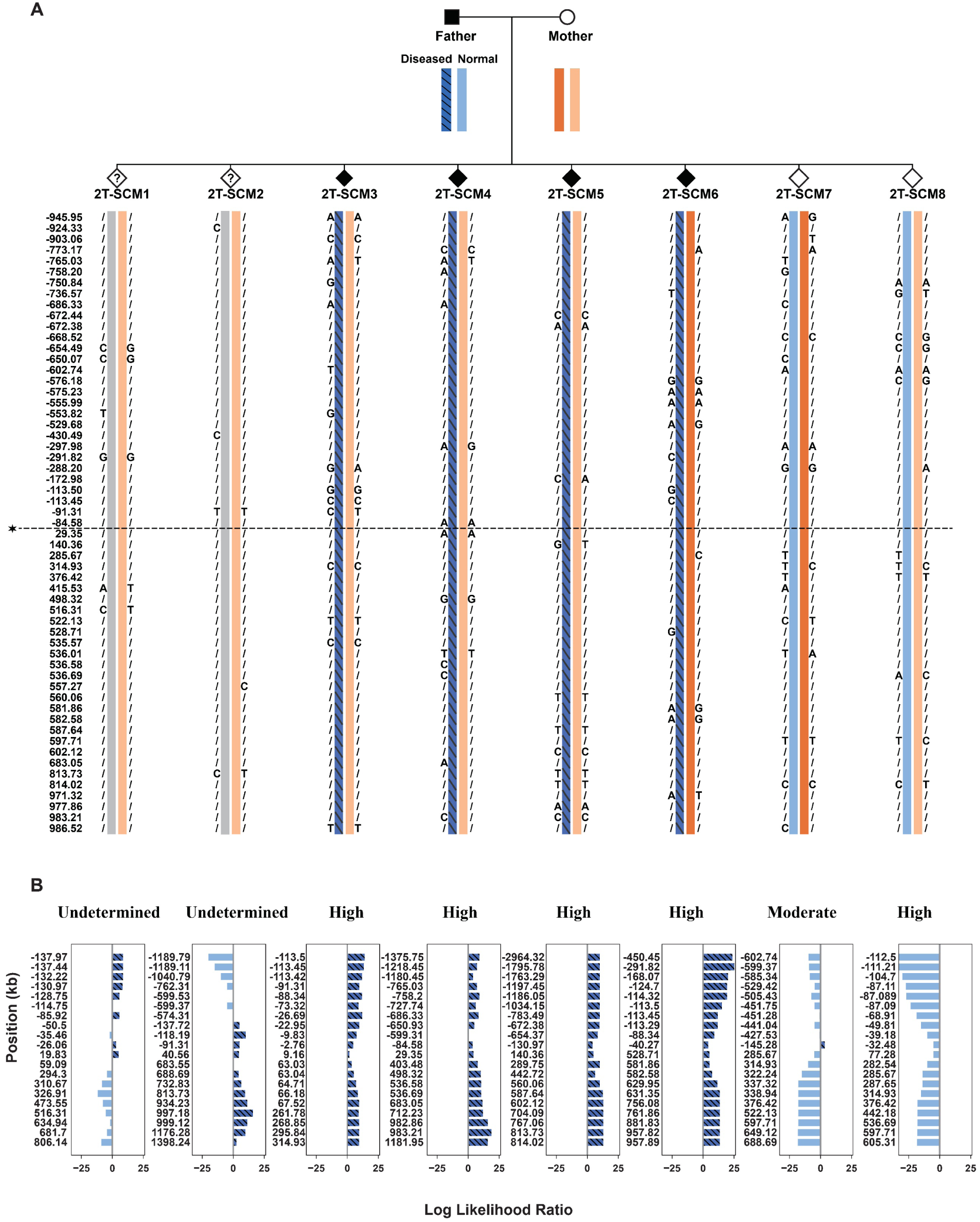
An illustrative case for niPGT-M. **A.** The SNP distribution and linkage analysis from direct cell-free DNA data in a single illustrative case in which the father (blue chromosomes) carried a genetic variant for Marfan syndrome (diagonal hatching, filled pedigree symbols), an autosomal dominant connective tissue disorder. Eight SCM samples were collected. **B.** For each embryo, numbers on y-axis indicate the chromosomal position centered on the causative locus (left). Log-likelihood ratio is plotted on the x-axis. Confidence in diagnosis is indicated above each graphic. Two samples were classified as ’Undetermined’ due to either a very low density of SNPs or a change in inference of allele inheritance very near the disease-carrying mutation site. All diagnoses were consistent with trophectoderm biopsies.

### Validation of niPGT-M using Matched Biopsies or Whole Embryos

We compared the results obtained from all SCM samples with results obtained from trophectoderm biopsy, discarded embryos, or amniotic fluid samples collected at two independent clinical centers (Table 1). We categorized the cases into three groups: autosomal dominant, autosomal recessive, and X-linked recessive disease. For autosomal recessive diseases, we assessed each embryo allele.

**Table 1.**
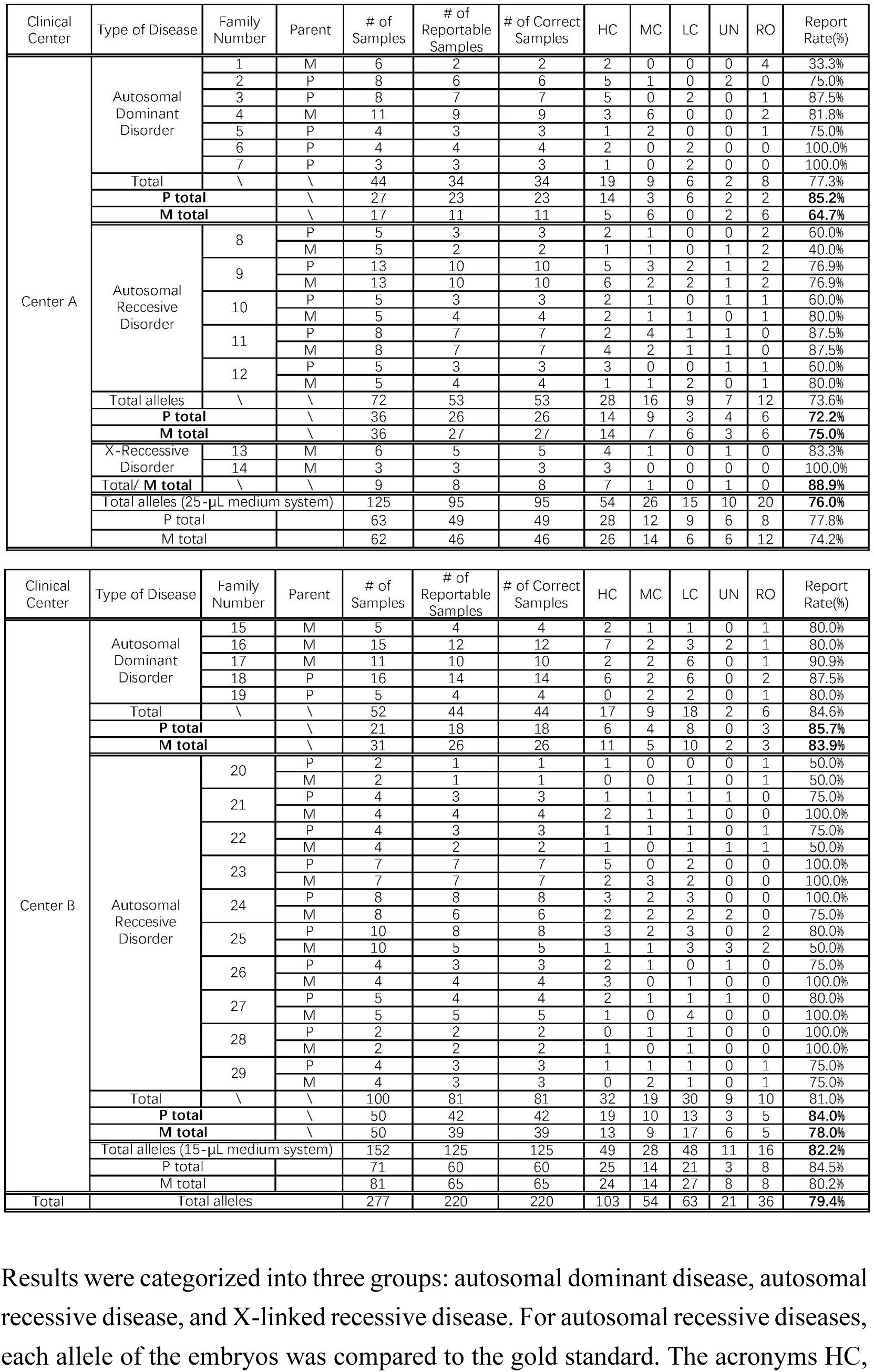
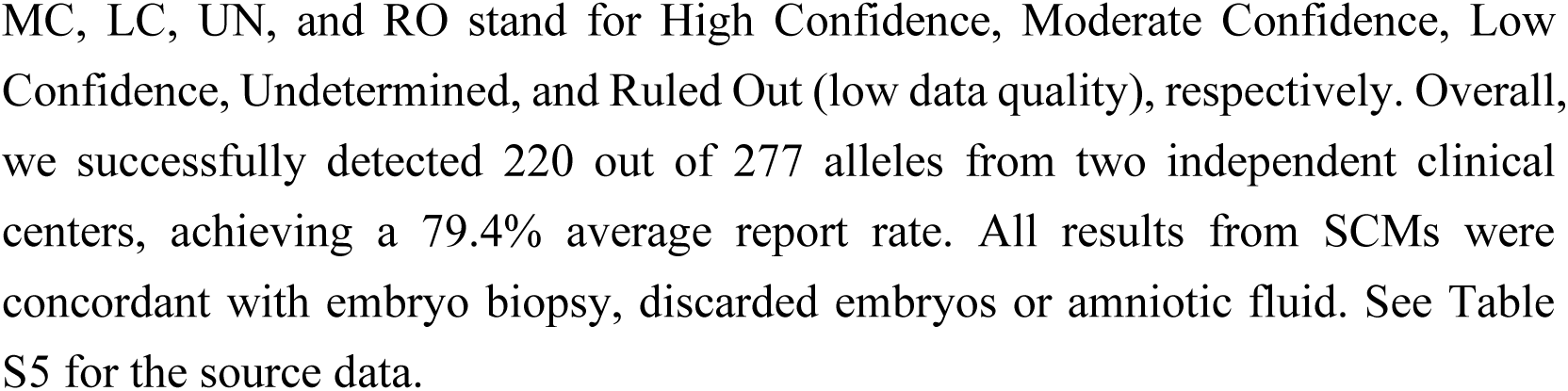
Summarized results of non-invasive preimplantation genetic testing for monogenic disorders using all matched biopsy/discarded embryos.

Samples from clinical center A were cultured using a 25-µL culture medium system (Methods). In the seven cases with autosomal dominant diseases, there were 44 SCM samples. For cases in which the father carried the causal mutation, 23 out of 27 SCM samples were successfully diagnosed (85.2% report rate). For cases in which the mother carried the causal mutation, 11 out of 17 SCM samples were successfully diagnosed (64.7% report rate). In the five cases with autosomal recessive disorders, 36 SCMs were analyzed. We successfully diagnosed the paternal allele in 26 samples (72.2% report rate) and the maternal allele in 27 samples (75% report rate). For the two cases with X-linked recessive disorders, 8 out of 9 samples (88.9% report rate) were diagnosed (5 male samples, 4 female samples; 1 male not reporting). Overall, we successfully diagnosed 95 out of 125 alleles derived from SCM samples from Clinical Center A (76.0% report rate). The overall report rate for paternal alleles was 77.8%, while the overall report rate for maternal alleles was 74.2%.

Independent validation was also performed in Clinical Center B, which used a 15 µL culture medium system (Methods). The five cases with autosomal dominant diseases, generated 52 SCM samples. Among the 21 SCM samples in which the father carried the causal mutation, 18 were successfully diagnosed (85.7% report rate). For the remaining 31 SCM samples in which the mother carried the causal mutation, 26 were successfully diagnosed (83.9% report rate). In the ten cases with autosomal recessive disorders, 50 SCMs were analyzed. We successfully diagnosed the paternal allele in 42 samples (84.0% report rate) and the maternal allele in 39 samples (78% report rate). Overall from Clinical Center B, we successfully diagnosed 125 out of 152 alleles from the SCM samples (82.2% report rate). The overall report rate for paternal alleles was 84.5%, while the overall report rate for maternal alleles was 80.2%.

The most important clinical criterion for evaluating preimplantation genetic testing for monogenic disease is the misdiagnosis rate, which is typically below 0.1% with conventional invasive methods, according to the ESHRE PGT consortium [27]. In all 220 reportable samples in our study, the diagnostic results were concordant with trophectoderm biopsy, discarded embryo, or amniotic fluid samples, indicating a diagnostic accuracy comparable to that of current invasive PGT methods (Table 1).

### Reconstruction of Embryo Genomes from Spent Embryo Culture Medium Data

The low amount of DNA released from blastocyst cells into the SCM limits the amount of genetic information that can be obtained directly. We therefore developed a Pedigree-Population-based Imputation method with Haploid Assumption to reconstruct the genomes of embryos (Fig. 3A and Methods).

**Figure 3.**
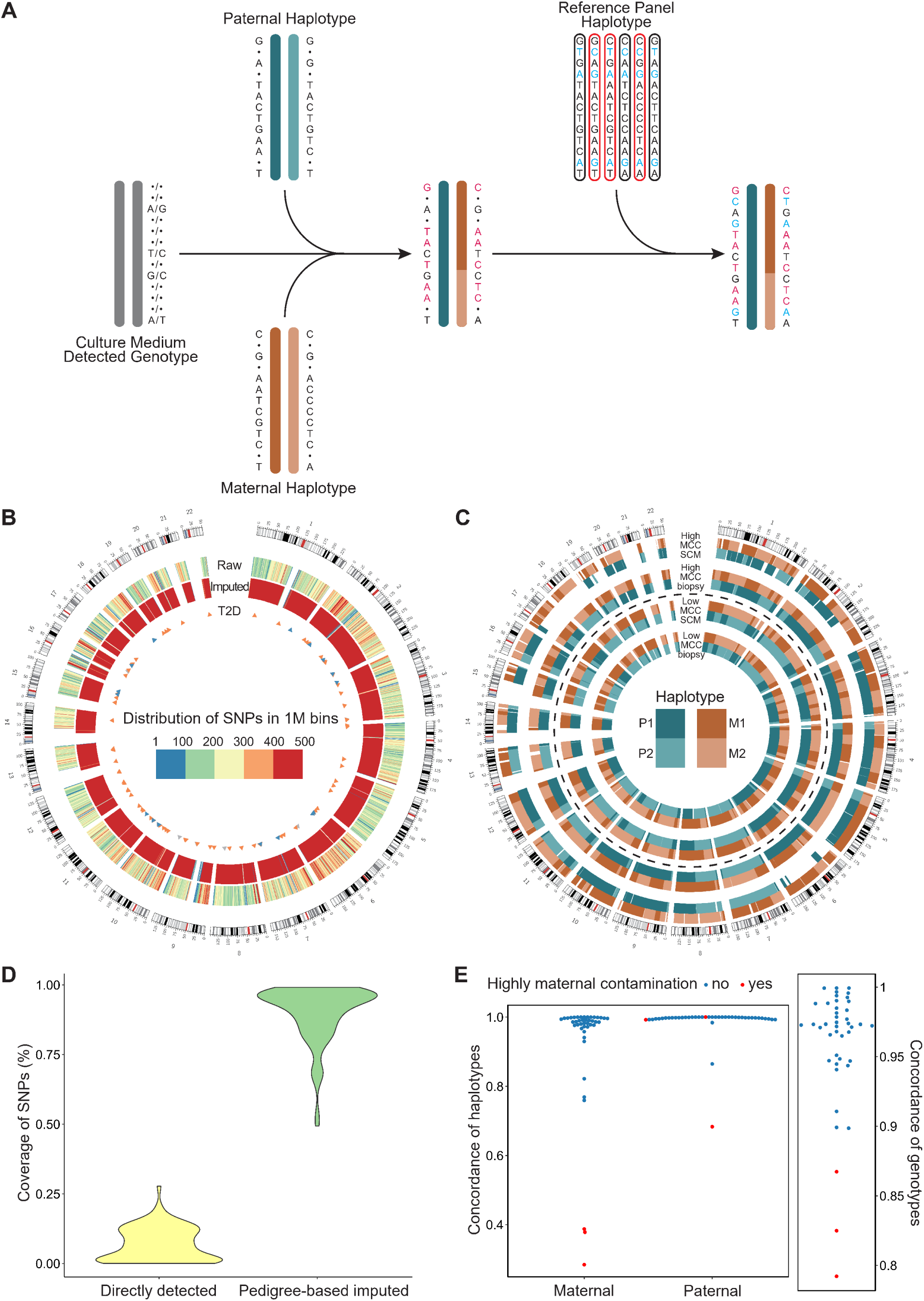
Reconstruction of embryo genomes from spent culture medium data. **A.** Schematic illustration of the procedure for genome reconstruction. This involves three steps. First, data was obtained from SCM (far left). Second, using the pre-phased parental haplotypes (blue and orange chromosomes) and selected informative SNPs that were detected directly, imputation was conducted with parental genome information, allowing creation of a haplotype scaffold of the embryonic genome. Third, embryo genomes were fully reconstructed using population-based genotype imputation. **B.** Whole-genome SNP density and Type II-Diabetes-related SNPs coverage in a SCM sample (10T-SCM1). Blue triangles indicate SNPs in the polygenic risk score model of Type II Diabetes directly called from raw data. Orange triangles indicate SNPs imputed from pedigree information. Gray triangles represent SNPs imputed from population information. SNP density is shown for raw and imputed data in 1 Megabase (M) bins. **C.** Haplotypes of two reconstructed embryonic genomes from SCM samples and their corresponding trophectoderm biopsy samples. One of the samples exhibited high MCC in the SCM (outer two rings) and the other exhibited low levels of MCC in the SCM (inner two rings). Maternal and Paternal haplotypes are indicated by color, with uncolored areas representing regions where reconstruction failed. **D.** SNP coverage of sequenced SCM samples from Clinical Center A before and after reconstruction. See Table S6 for source data. **E.** Haplotype and genotype concordances between the reconstructed embryonic genome from SCM samples collected at Clinical Center A and that from the corresponding trophectoderm biopsy, discarded embryos or amniotic fluid samples. See Table S6 for the source data. There is no quantitative x-axis on panels D and E; points are spread horizontally for visualization.

The initial median density of detected SNPs in the raw data from Clinical Center A was 222 SNPs per megabase (Mb)(IQR, 161 to 271 SNPs/Mb, Fig. 3B). After performing reconstruction (Fig. 3A, Extended Data Fig. 8), the median density of imputed SNPs in data derived from SCM increased to 1514 SNPs per Mb (IQR, 1196 to 1805 SNPs/Mb, Fig. 3B), with the coverage of SNPs detected in embryo genomes from SCM improving from a median of 6.4% (IQR, 1.2% to 11.8%) to a median of 94.0% (IQR, 86.5% to 96.7%) (Fig. 3D).

We compared the predicted embryo’s haplotype with the haplotype of corresponding discarded embryos or trophectoderm biopsy samples. When the MCC level was low, we observed that the haplotype configurations of SCM and the corresponding gold standard were highly similar (Fig. 3C, inner rings). Conversely, SCM samples with high levels of MCC exhibited lower haplotype similarity with gold standards, especially in maternal haplotypes (Fig. 3C, outer rings). For further exploration, we obtained samples with paired genome sequencing data from both SCMs and their corresponding discarded embryos or trophectoderm biopsy samples, and removed from further analysis samples exhibiting aneuploidy or poor quality-control metrics (Methods, Table S4). Paternal or maternal haplotype concordance at each physical site was used to measure the accuracy of reconstructed haplotype (Fig. 3E). For samples without significant MCC, we found that the median concordance of maternal haplotypes in SCM was 98.6% (IQR, 97.2%-99.7%), while the median concordance with paternal haplotypes was 99.8% (IQR, 99.5% to 100%) (Fig. 3E). High levels of MCC affect haplotype reconstruction from SCM, especially maternal haplotypes (Fig. 3E).

We next compared the genome-wide genotypes reconstructed from SCM to the corresponding gold standard (trophectoderm biopsy or discarded embryos) genotypes. The concordance of genotypes ranged from 79.2% to 99.9%, and, consistent with the previous analysis, the three samples with the lowest concordance (below ∼0.9) suffered from high MCC (Fig. 3E). For those samples without significant MCC, the median genotype accuracy was 97.3% (IQR, 96.2% to 98.8%). We also compared the reconstructed genotypes from two SCM samples with the genotypes from corresponding amniotic fluid samples, and observed concordance rates of 94.5% and 94.1%. Prior to all comparisons, we removed SNPs showing Mendelian errors in gold standard samples, which never exceeded 1% of all SNPs.

### Polygenic Risk Evaluation of Type II Diabetes for Embryos

We used Type II Diabetes as a model complex disease to assess the feasibility of polygenic risk evaluation based on genomes reconstructed from SCM data collected from Clinical Center A. We first applied data quality filters to remove samples with high MCC, chromosomal abnormalities, or other metrics indicating poor quality (Supplementary Methods, Table S4). This left 25 samples for calculation of polygenic risk score using a model for Type II Diabetes from a previous study [39]. The model involves a total of 114 loci. In the 25 samples selected for this analysis, 28.7% of the loci were directly genotyped, 52.5% were inferred through pedigree-based genome reconstruction, and the remaining 18.8% were imputed via population-based reconstruction (Fig. 3B, Fig. 4A).

**Figure 4.**
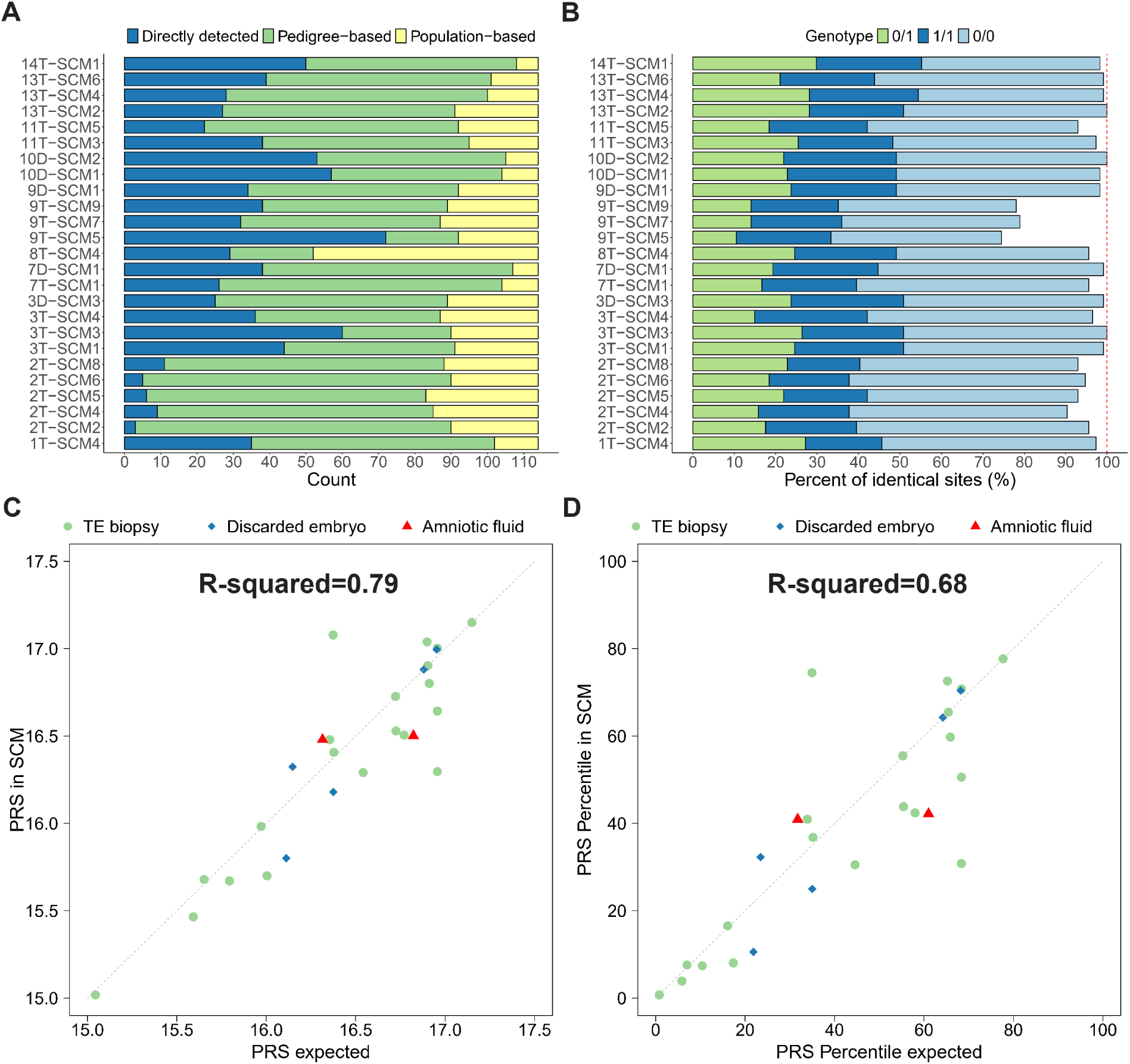
Type II Diabetes associated genotype imputation and polygenic risk score calculation. **A.** Distribution of reconstructed genotypes of Type II Diabetes-associated SNPs in select SCM samples from Clinical Center A. Type II Diabetes related genotypes were obtained from direct detection by next-generation sequencing in cell-free DNA from SCMs (blue bar), pedigree-based genotype imputation (green bar) and population-based genotype imputation (yellow bar). **B.** Genotype concordance of Type II Diabetes-associated SNPs in SCMs versus corresponding trophectoderm or discarded embryo samples. Genotypes 0/0, 1/1, and 0/1 respectively denote homozygous reference allele, homozygous alternate allele, and heterozygous genotypes. **C.** Consistency of Type II Diabetes polygenic risk score (PRS) between SCM and paired trophectoderm (light green circles), discarded embryo (blue diamonds), or amniotic fluid (red triangle) samples. **D.** Polygenic risk score percentile of Type II Diabetes between SCM and paired trophectoderm (light green circles), discarded embryo (blue diamonds), or amniotic fluid (red triangle) samples. Trophectoderm samples were additionally available for two amniotic fluid samples. See Table S7 for the source data.

When comparing the imputed genotypes of each embryo derived from the SCM samples to the genotype derived from the corresponding discarded embryos or trophectoderm biopsy samples, 94.6% of the SNPs were identical (average among embryos, Fig. 4B). Among the identical SNPs, 52.8% are homozygous reference (hg19) genotypes, 24.9% are homozygous alternate genotypes, and 22.3% are heterozygous (Fig. 4B).

The polygenic risk score of Type II Diabetes calculated from SCM samples showed a high level of concordance with those calculated from the corresponding discarded embryos, trophectoderm biopsy samples, or amniotic fluid, with an R-squared value of 0.79 (Fig. 4C). In clinical practice, polygenic risk score percentiles are typically used to evaluate the relative genetic risk of common diseases instead of raw polygenic risk scores. Generally, the higher the percentile, the greater the risk of a genetic disease. We observed a correlation in polygenic risk score percentiles derived from SCM samples and their corresponding discarded embryos, trophectoderm biopsy samples, or amniotic fluid samples (R-squared value of 0.68, Fig. 4D).

## Discussion

We overcame challenges posed by DNA-fragment length amplification biases and high albumin concentration in samples by reengineering the LIANTI method to amplify trace DNA from SCM. To diagnose monogenic disorders, we developed a linkage analysis method that accounts for high allele drop-out and maternal DNA contamination. This method detects disease-carrying chromosomes with extremely low false-positive and false-negative rates and assigns a confidence level to each diagnosis. The three confidence levels are derived from posterior probabilities, and the underlying log-likelihood ratio curve can be examined if a more nuanced confidence estimate is desired. Confidence ratings were intentionally designed to be highly conservative, which is reflected empirically in the accuracy (220/220) observed across all reported samples. Our approach addresses the central challenges in using SCM for genetic assessments, achieving diagnostic-grade accuracy among reportable samples, substantially reducing the risks associated with embryo biopsy, and laying a technological foundation for clinical adoption.

To further assess the robustness and generalizability of our approach, we performed an independent validation in a second clinical center operating under a different embryo culture volume and the same workflow. Across 15 additional families, our method achieved a report rate of 82.2%, slightly higher than that observed in the original center, while maintaining 100% concordance with gold-standard results for all reportable samples. This indicates that the accuracy and reporting characteristics of our niPGT framework are reproducible across clinical settings. The consistency in report rate further suggests that reporting efficiency may be amenable to technical optimization.

Broad clinical adoption will require further improvement in report rate and workflow integration. Nonetheless, even at the present stage, there are specific clinical contexts where our niPGT protocol may provide value. In scenarios where biopsy poses heightened concern, such as when embryo quality is compromised or when only a small number of embryos is available, clinicians and patients often face a difficult balance between obtaining genetic information and avoiding potential harm. In such circumstances, non-invasive PGT offers an additional, low-risk option. Any reliable diagnostic information obtained from the embryos (even if only from a large fraction) may assist in decision-making without introducing procedural risk to the embryo. niPGT is also an option for parents who are psychologically or ethically reluctant to subject their embryos to invasive procedures [40], thereby enabling genetic testing that would otherwise be avoided entirely.

Technical advances are likely to further broaden the applicability of niPGT. If non-invasive testing can be accelerated to deliver results within hours, it may become feasible to perform niPGT prior to cryopreservation. In such cases, embryos with non-reportable niPGT results could still undergo subsequent biopsy if needed, thereby combining the safety advantages of non-invasive testing with the advantages of existing clinical workflows. Declining sequencing costs and increased automation will result in cost parity between niPGT and traditional invasive PGT, if it has not been achieved already. Particularly important is that niPGT offers a clear path toward scalable, standardized, and operator-independent workflows. By minimizing dependence on highly skilled clinicians, niPGT can simultaneously improve procedural consistency and reduce personnel costs. Finally, maternal DNA contamination from cumulus cells or polar bodies in SCM remains a critical challenge affecting both niPGT-M and niPGT-P. Developing improved protocols to prevent MCC or remove maternal DNA should therefore be a priority. Until such protocol improvements, the simple method for detection and quantitative estimation of MCC levels that we developed here can be used to prevent misinterpretation of results when maternal contamination is high, and provide increased confidence in results when MCC is low.

In contrast to trophectoderm samples that nearly always contain alleles from both haplotypes, SCM has a high rate of allelic drop-out, resulting in single-haplotype segments in the sequencing data. To address this issue, we developed a specialized approach to estimate the haplotype status of each SNP. Haplotype status in this case is defined as whether both haplotypes, only the paternal haplotype, or only the maternal haplotype was detected. In modeling SNP haplotype status, an assumption of independence among measured SNPs would introduce an excessive number of parameters, leading to the risk of overfitting. To account for this, we adopted a cautious approach in which the default assumption is that both parental haplotypes are detected unless the SNP data strongly indicates otherwise.

Related to the issue of allelic dropout, our current niPGT-M approach relies on linkage analysis because direct interrogation of the exact pathogenic site is generally not feasible, given the trace and fragmented DNA present in spent culture medium. Linkage-based diagnosis is a validated and robust strategy in clinical PGT-M, provided that accurate parental haplotype phasing is achieved. While our Bayesian linkage analysis builds upon classical HMM-based haplotyping logic, it has been specifically adapted to model the high allele drop-out and maternal contamination characteristic of cfDNA from spent culture medium, enabling robust inference where conventional algorithms fail. In cases where affected relatives are unavailable, sequencing data from discarded embryos must be used for pre-phasing. However, the data quality and coverage obtained from discarded embryos are often limited, which can hinder the accuracy of haplotype phasing results for the parents. Recent advances in long-read and single-cell sequencing technologies may now enable accurate parental haplotype phasing using only parental DNA [41,42], eliminating the need for additional family members and further enhancing the applicability of niPGT-M. Finally, de novo mutations remain challenging for linkage-based inference. However, this situation represents a very small fraction of PGT-M cases, as most clinical PGT-M indications involve families with known inherited pathogenic variants.

Studies have shown that embryo selection based on PGT-P can result in substantial and clinically relevant reductions in disease risk [31]. At the same time, weighty ethical concerns surrounding PGT for polygenic disorders have led many countries to ban the practice in the context of IVF. The non-invasive approach presented here was aimed primarily at reconstructing embryo genomes and accurately estimating polygenic risk from SCM. By achieving this goal, this work establishes a safer and technically validated protocol for genome-wide embryo genotyping, enabling further research into the feasibility and implications of polygenic risk assessment. Clinical application of this technology must proceed hand-in-hand with rigorous ethical review and broad societal dialogue.

Although we achieved 100% accuracy (220/220) in all reported samples across multiple embryos and monogenic disease contexts, diagnostic errors are inevitable in larger cohorts. Nonetheless, the results suggest that the error rate of the niPGT method presented here will be very low in reportable samples. Importantly, a confidence classification is provided for each diagnosis, helping clinicians and parents make more informed decisions about the most suitable course of action. The method’s accuracy, interpretability, and compatibility with standard IVF workflows together establish a strong foundation for translation of niPGT to the clinic.

## Methods

### Institutional Review Board Approval

This study was approved by the Peking University Third Hospital Ethical Review Committee (Approval no. 2020SZ-005) and the Ethics Committee of Chinese PLA General Hospital (Approval no. S2022-586-01). The patients/participants provided written informed consent to participate in this study.

### Study Subjects

Included subjects (n=29 couples) were using intracytoplasmic sperm injection and PGT with trophectoderm biopsy for preventing transmission of inherited genetic diseases at Peking University Third Hospital (Clinical Center A) between September 2019 to April 2021 and at First Medical Center of PLA General Hospital between September 2022 to November 2024 (Clinical Center B). All 29 of the patients had genetic testing reports.

### Assisted Reproductive Technology (ART) Laboratory Protocols and Blastocyst Biopsy for PGT-M

Standard protocols were used for intracytoplasmic sperm injection [43]. The zygotes were cultured in equilibrated Vitrolife G-series media with human serum albumin (HSA; LifeGlobal), overlain with mineral oil in incubators with 6% CO_2_ and 5% O_2_ at 37°C until day 3. On day 3, embryos were moved to and cultured individually in fresh 25-μL (Clinical Center A) or 15-μL (Clinical Center B) microdrops of G-series media with HSA. Embryos were evaluated on day 5, 6, or 7 for trophectoderm (TE) biopsy. A few TE cells from each hatching blastocyst were biopsied and sent to the PGT lab for PGT-M testing.

### Collection of Culture Medium Samples and Discarded Embryos

Immediately after the blastocysts were transferred to another dish for biopsy, 20 μL (25 μL culture system) or 13 μL (15 μL culture system) of SCM was removed from each of the residual spent medium drops. For discarded embryos, each embryo was gently moved with a pipette tip to the edge of its microdrop and then removed and transferred into a RNase-DNase-free PCR tube containing 3 μL of lysis buffer. The corresponding SCM was collected for analysis. Pipette tips were changed between collections of each sample to avoid cross-sample contamination. Media drops that were incubated and collected under identical conditions to those used for blastocyst culture but that never contained embryos served as negative controls. All samples were immediately frozen after collection and stored at -80°C until analyzed.

### Spent Embryo Culture Medium Treatment and Discarded Embryo Lysis

Frozen 20-μL and 13-μL SCM samples were thawed and gently mixed, and 10.8 μL was removed to a PCR tube (Maximum Recovery, Axygen) containing 1.2 μL 10x lysis buffer (1x lysis buffer: 60 mM Tris-Ac pH 8.3, 2 mM EDTA pH 8.0, 15 mM DTT, 0.5 μM carrier ssDNA (5’-TCAGGTTTTCCTGAA-3’, Thermo Fisher Scientific oligo with PAGE purification)), briefly centrifuged, then heated at 75°C for 30 min. 0.5 μL 35mg/mL QIAGEN protease (dissolved in water and stored at 4°C) was added and incubated at 55°C for 2 hrs followed by 80°C for 30 min. The resulting lysate in PCR tubes was immediately subjected to the reengineered linear transposon-based amplification as described herein or stored at -20°C for later use. Frozen discarded embryos were thawed and heated at 75°C for 30 min, followed by addition of 0.5 μL 5mg/mL Qiagen Protease and heating (2 h at 55°C and 30 min at 80°C) in 3 μL of lysis buffer.

### Whole Genome Amplification

Transposon DNA (5’/Phos/CTGTCTCTTATACACATCTGAACAGAATTTAATACGACTCACTATAGGGAGATGTGTATAAGAGACAG-3’, Thermo Fisher Scientific oligo with PAGE purification) was annealed into self-looping structure by gradual cooling in annealing buffer (20 mM Tris-Ac pH 8.3, 50 mM NaCl, 2 mM EDTA pH 8.0) with a final DNA molecule concentration of a 1.5 μM. The 1.5 μM annealed transposon DNA was then mixed with an equal volume of ∼1 μM Tn5 transposase (Lucigen, EZ-Tn5™ Transposase) and incubated at room temperature for 30 min, dimerizing into transposomes with a final concentration of ∼0.25 μM. The transposomes can be stored at -20°C. Each single-cell transposon-based amplification requires 0.5 μL of this transposome solution.

Starting with the culture medium lysate, a transposition mixture was assembled by addition of MgCl_2_ to 2.5 mM and transposome to 18.75 nM in a total volume of 20 μL. The transposition reaction was carried out at 55°C for 12 min. A 0.84 μL mixture of EDTA (200mM, 0.5ul), NaCl (5M, 0.17ul), and ssDNA (50uM, 0.17ul, 5’-TCAGGTTTTCCTGAA-3’) was then added to each tube, followed by incubation at 68°C for 30 min. For end-filling and extension, A 1.6 µL mixture containing 1 µL of 100 mM MgCl₂, 0.4 µL of 10 mM dNTP, and 0.2 µL of 1 M DTT was added to each tube from the previous step. After thorough mixing, 0.2 μL Q5 HF DNA Polymerase (New England Biolabs) was added and the reaction mixture was heated at 73°C for 45 seconds. 0.5 μL 4mg/mL Qiagen protease was added and the PCR tube was heated at 50°C for 1 hour in the presence of 4 mM EDTA to inactivate DNA polymerase, followed by 2.33 µL of 5 M NaCl to achieve a 450 mM NaCl concentration in a total reaction volume of ∼25 µL. Protease heat inactivation was performed by incubation at 77°C for 20 minutes. DNA fragments were amplified to RNAs in the same tube by adding a 63.8ul T7 in vitro transcription reaction mixture (34.7ul for H2O, 0.7ul for 50 uM ssDNA(5 ’ - TCAGGTTTTCCTGAA-3’), 9ul for 10X RNAPolymease Buffer, 0.9ul for 1M DTT, 6.5ul for 25mM each rNTP Mix, 9ul for 100% DMSO, 3ul for 3.2 U/μL SUPERase In RNase Inhibitor) overnight at 37°C as described in Chen et al [22].

The next day, 10 μL 0.5M EDTA was added to each tube and RNAs were column purified (Zymo Research). 18 μL RNAs were transferred to a PCR tube containing a 3.1 μL solution of 6.5mM each dNTPs, 9.7 μM carrier ssDNA (5’-TCAGGTTTTCCTGAA-3’, Thermo Fisher Scientific oligo with PAGE purification) and 3.2 U/μL SUPERase In RNase Inhibitor (Invitrogen). The denaturation incubation was at 70℃ 1 min, 90℃ 15s, and followed by ice quenching. The reverse transcription reaction for the first strand was carried out in a 30 μL SuperScript IV reserve reaction system including 0.67 mM each dNTP, 0.6 U/μL SUPERase In RNase Inhibitor (Invitrogen) and 6 U/μL SuperScript IV Reserve Transcriptase (Invitrogen) in SuperScript IV buffer. The sequence of the primer was 5’-AGATGTGTATAAGAGACAG-3’ (Thermo Fisher Scientific oligo with PAGE purification), and the incubation program was 25°C 1 min, 37°C 1 min, 42°C 1 min, 50°C 1 min, 55°C 15 min, 60°C 10 min, 65°C 12 min, 70°C 8 min, 75°C 5 min, 80°C 10 min. RNA was then removed by incubation at 37°C for 30 min with 10 ng/μL affinity-purified RNase A (Invitrogen) and 0.08 U/μL RNase H (New England Biolabs). Second strand synthesis was carried out in a 100 μL Q5 DNA polymerase system (New England Biolabs, 1X Q5 reaction buffer, 1X Q5 High GC enhancer, 200 μM dNTPs, 0.5 μM primer, 0.02 U/μL Q5 DNA polymerase), with the primer 5’-NNNNNNNNGGGAGATGTGTATAAGAGACAG-3’ (Thermo Fisher Scientific oligo with PAGE purification). Each tube was heated at 98°C for 30 s, then 10 cycles of 10 s at 98°C, 30s at 58°C, 30s at 60°C, 30s at 65°C, 2.5 min at 70°C, then 72°C 6 min for strand extension. The resulting amplicons were column purified into 23 μL elution buffer and stored at -20°C. The DNA concentration of the product after amplification was measured using a Qubit 2.0 fluorometer (ThermoFisher Scientific) with the Qubit dsDNA HS Assay kit (Life Technologies).

Starting with the discarded embryo lysate, DNA was amplified following the LIANTI amplification method described in [22], except that the sequence of the DNA primer named 19_1 was (5’-AGATGTGTATAAGAGACAG-3’). Primer 19_1 was added to the first-strand cDNA synthesis reaction.

### Sequencing and Data Analysis

For library preparation, the nucleic acid amplification product was sonicated (Covaris S2) to the length required by each sequencing platform. For 2×150 bp paired-end Illumina sequencing, library preparation was performed by NEBNext Ultra II DNA Library Prep Kit for Illumina (New England Biolabs), following the instructions of the manufacturer and skipping the optional size selection step. Library preparation was performed by a PCR-free Library Prep Kit, NEBNext Ultra II DNA Library Prep Kit for Illumina (NEB #E7645S/L). Genomic DNA (gDNA) samples, SCM samples, and discarded embryo samples were sequenced on the Illumina NovaSeq 6000 System or Illumina HiSeq 4000 platform with 2×150 bp pair-end sequencing. Sequencing lanes were shared by 24 samples with NEBNext Indexes (NEB #E6609S/L). Each gDNA sample was sequenced to generate ∼90 Gb raw data and each SCM sample or discarded embryo was sequenced to generate ∼30 Gb raw data.

### Mapping and SNP Calling

Raw paired-end reads were trimmed of adapters and low-quality ends by cutadapt [44]. Clean reads were mapped to human reference genome hg19 with BWA mem [45]. Each BAM file was screened for duplicates and sorted by MarkDuplicatesSpark of GATK 4.2.0 [46]. Base quality score recalibration (BQSR) was performed using BaseRecalibrator to generate a recalibration table, followed by ApplyBQSR to adjust the base quality accordingly. For variant calling, GATK HaplotypeCaller produced a genomic variant call format (GVCF) file for each sample. GenotypeGVCFs was utilized to perform joint genotyping across all samples. SNPs were filtered using variant quality score recalibration (VQSR) with HapMap 3.3, Omni 2.5, 1000 Genome phase I, and dbSNP build 151 as SNP training sets. A 99% sensitivity threshold was chosen to filter SNPs accurately. For subsequent analysis, we exclusively retained SNPs for which only two different alleles are observed in the population. In the pre-phasing stage, to deduce the two parental haplotypes, we employed Plink 1.9 [47] with the option “--mendel” to identify sites with Mendelian errors.

### Parental haplotype phasing

For parental haplotype phasing, genetic information from sibling embryos or other family members is needed. A likelihood-based haplotyping approach using a Hidden Markov Model (HMM) and dynamic programming Viterbi algorithm was used to determine the most likely haplotype configuration of parents [37,38]. The transition probability of haplotype state changes between consecutive loci was calculated from the recombination fraction obtained from the genetic distance map of the 1000 Genomes Project Phase 3 references (https://www.internationalgenome.org/). To address the allele dropout issue stemming from scWGA bias during parental haplotyping with sibling embryos, we filtered out sites that violated Mendelian rules or phased variants that did not adhere to chromosome interference theory [48]. Each chromosome of each parent was then independently processed until the parental haplotypes of all chromosomes were phased.

### BASE-niPGT-M: a BAyesian linkage analySis mEthod for niGPT-M

We developed a computational Bayesian linkage analysis method for niPGT-M (Extended Data Fig. 7 and Supplementary Methods). The model used sequencing data from SCM and the phased haplotypes of the parents as input. Our approach incorporates the fraction of maternal cell contamination and haplotype status of each SNP into the likelihood function.

Both entire samples and individual SNPs were assessed for data quality, with low-quality samples and SNPs eliminated. For each sample, the sequencing error rate, fraction of maternal cell contamination and haplotype status were estimated. The fraction of maternal cell contamination was defined as the fraction of DNA fragments originating from the maternal chromosome among all cell-free DNA in the solution. Haplotype status, representing the true parental origin of DNA at each measured SNP, is categorized into three distinct classes: ’Paternal Chromosome Only,’ ’Maternal Chromosome Only,’ or ’Both Parental Chromosomes’. For instance, the designation ’Paternal Chromosome Only’ indicates that the DNA identified at a specific SNP exclusively originates from the father. Initial fraction of maternal cell contamination and haplotype status were estimated, and these parameters were then iteratively and jointly calibrated until convergence (Supplementary Methods).

Starting from the disease-causing mutation site, SNPs were incrementally incorporated one by one, calculating the log-likelihood ratio of inheriting the pathogenic allele versus the corresponding reference allele for each SNP subset, ultimately resulting in a log-likelihood ratio curve (Fig. 1B). The calculation is based on the single-SNP likelihood, which was obtained from a binomial model using the measured allelic depth values and estimated parameters, and the recombination probability, which can be obtained from databases containing recombination fractions. Typically, the curve initiates at a point where the ordinate is in close proximity to zero, gradually stabilizing into a plateau with the addition of sufficient SNPs. This log-likelihood ratio curve enabled the identification of the inherited allele at the locus of interest, and categorized the diagnostic results for each sample into four classes: High Confidence, Moderate Confidence, Low Confidence, and Undetermined. The criteria for these categorizations were set with clinical guidance in mind, and are based on both probabilistic principles and conservative biological and technical considerations such as the integrity of haplotype blocks and possible sequencing errors. Practically, a High Confidence diagnosis is almost certain to be correct (Posterior probability >99.9%), while a Moderate classification corresponds to strong evidence (Posterior probability >95%) but with clear indicators of uncertainty. Low confidence diagnoses have some supporting evidence but should be treated with caution. See Extended data Fig. 7 and Supplementary Methods for more details.

### Haplotype phasing and whole genome reconstructing from SCM

We developed a Pedigree-Population-based Imputation with Haploid Assumption (PPIHA) method to reconstruct the embryonic genome from the sequencing data of SCM (Fig. 3A and Extended Data Fig. 8).

We first combined pre-phased parental haplotype information and selected informative SNPs (Extended Data Fig. 9) where at least one parental origin haplotype could be confidently phased to construct a sparse haplotype scaffold with a single parent. Paternal or maternal haplotypes in the scaffold were first corrected if continuous variants of the opposite haplotype were observed on both nearby sides. Then with a parental haplotype and Hidden Markov Model strategy, paternal and maternal haplotypes of other variants were independently phased based on the predetermined haplotype scaffold (Extended Data Fig. 9). Joint probabilities of the two states of paternal or maternal haplotypes at each variant were calculated using a forward-backward algorithm based on the adjoining nearby ten variants. The haplotype of the variant was then assigned to the state with the higher probability. By organizing the haplotype states of all variants according to their chromosomal positions, the pedigree-based haplotypes of SCMs were reconstructed. Subsequently, the genotypes of SCM were imputed with the genome information of the parents (Fig. 3A, Extended Data Fig. 9).

Following pedigree-based genome reconstruction, population-based genotype imputation was conducted using the 1000 Genomes haplotype reference panel with Minimac4 [49], resulting in the reconstruction of the embryonic genome (Supplementary Methods).

We then employed a polygenic risk score model for Type II Diabetes including 128 SNPs from a previous study [39]. SNPs whose genotype was not available or failed to be imputed in the UK Biobank cohort were excluded, leaving a total of 114 SNPs. A reference population from the UK Biobank was used to construct the polygenic risk score distribution for Type II Diabetes. Each embryo’s polygenic risk score as well as its percentile for Type II Diabetes was obtained, and compared with those calculated from trophectoderm biopsy, discarded embryo, or amniotic fluid samples as the gold standard.

## Data availability

Access to anonymized patient data is subject to a data-sharing agreement and protocol approval from the institutional review board committee. The study-specific analyzable dataset is, therefore, not publicly available. Requests for data should be directed to the corresponding authors. All summary statistics and source data are available in Supplementary Tables.

## Code availability

Code for BASE-niPGT-M is available at https://github.com/Ge-lab-pku/BASE-niPGT-M

## Acknowledgements

We thank Long Gao, Yuhang Huan, Ye Li, Cuiping Pan, Xiaodan Shi, Wenjie Sun, Zhongwei Wang, Liying Yan, Jingyi Yang, Jiankun Zhang and Nannan Zhang for their help. This work is funded by Beijing Advanced Innovation Center for Genomics at Peking University and Changping Laboratory. H.G. is supported by National Natural Science Foundation of China (No. T2225001). This study was also supported by the National Key R&D Program of China (2023YFC2705604), the National Science Foundation of China (82071721 and 82371706), the special fund of the National Clinical Key Specialty Construction Program, P. R. China (2023) and the National Key Research and Development Program of China (2024YFC2707100).

## Author contributions

XSX, JQ, LH, SL, HG and HP conceived the research. JH, JJ and MM collected SCM samples and performed PGT-M experiments of biopsy samples. LH designed and performed the niPGT-M experiments with help from GY, DC, YT and KL. RZ and HG performed the lineage analysis for niPGT-M with the help from LH, QW, ZW, WS and XC. YZ and YX performed the data analysis for niPGT-P with help from LH and GC. LH, HG, YX and YZ wrote the initial manuscript with help from QW and GY. All authors approved the final version of the manuscript.

## Additional Information

Extended data Figs.

Supplementary Methods

Supplementary Tables

**Extended Data Fig. 1.**
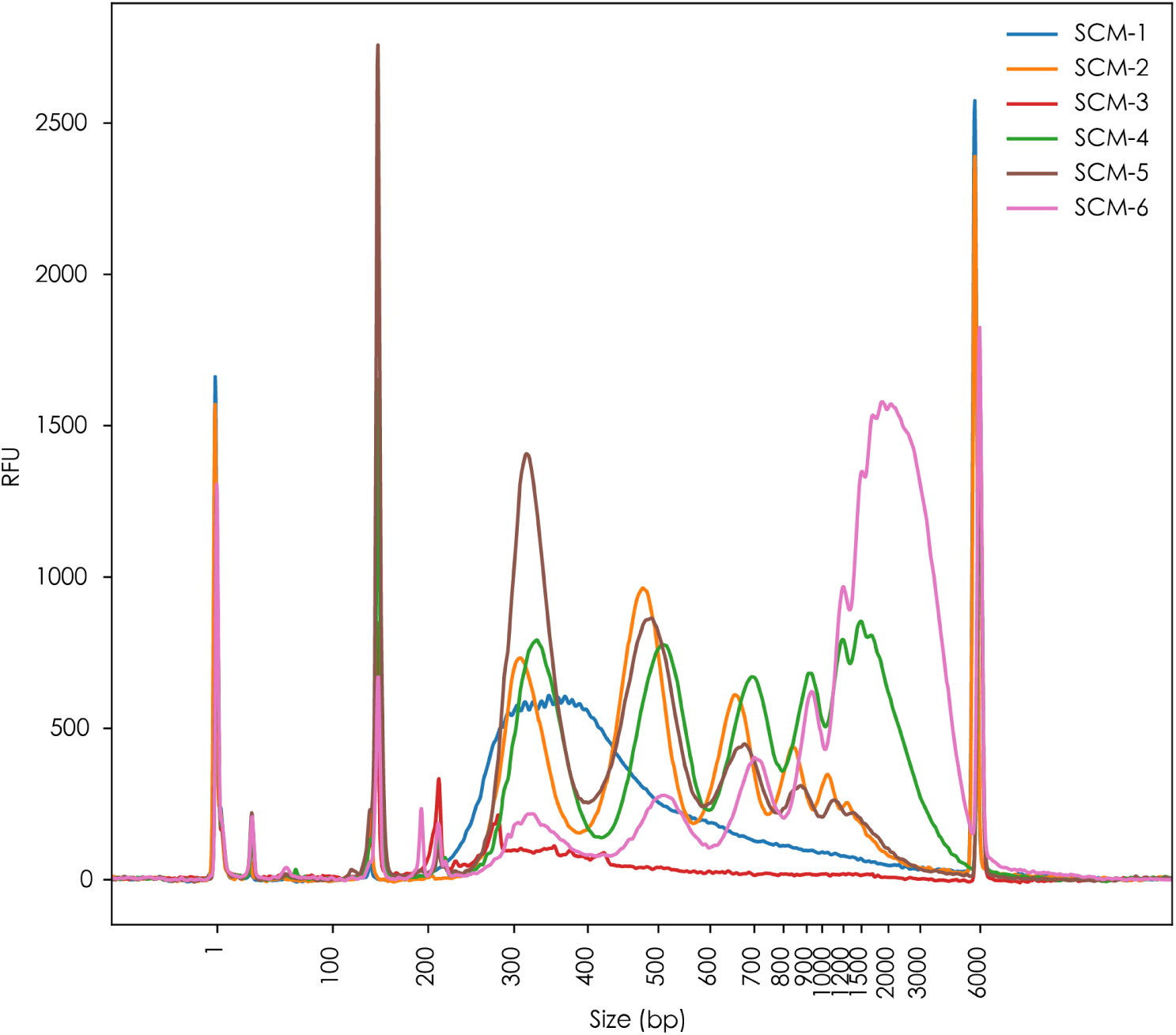
DNA length distribution in spent embryo culture medium. Length distributions of DNA molecules in six spent embryo culture medium samples (Agilent Fragment Analyzer System). The horizontal axis represents the length of DNA fragments detected from 1 to 6,000 bp. The vertical axis represents a unit of relative fluorescence units. The first peak of insert fragments was approximately 180 bp, with subsequent peaks spaced at approximately 180 bp intervals, corresponding to the length of DNA wrapped around a nucleosome. Samples SCM 2, SCM4, SCM5 and SCM6 showed varying levels of apoptosis, suggesting different degrees of DNA degradation. SCM3 failed to produce an effective library due to its low cfDNA content. SCM1 did not exhibit the characteristic apoptosis signals.

**Extended Data Fig. 2.**
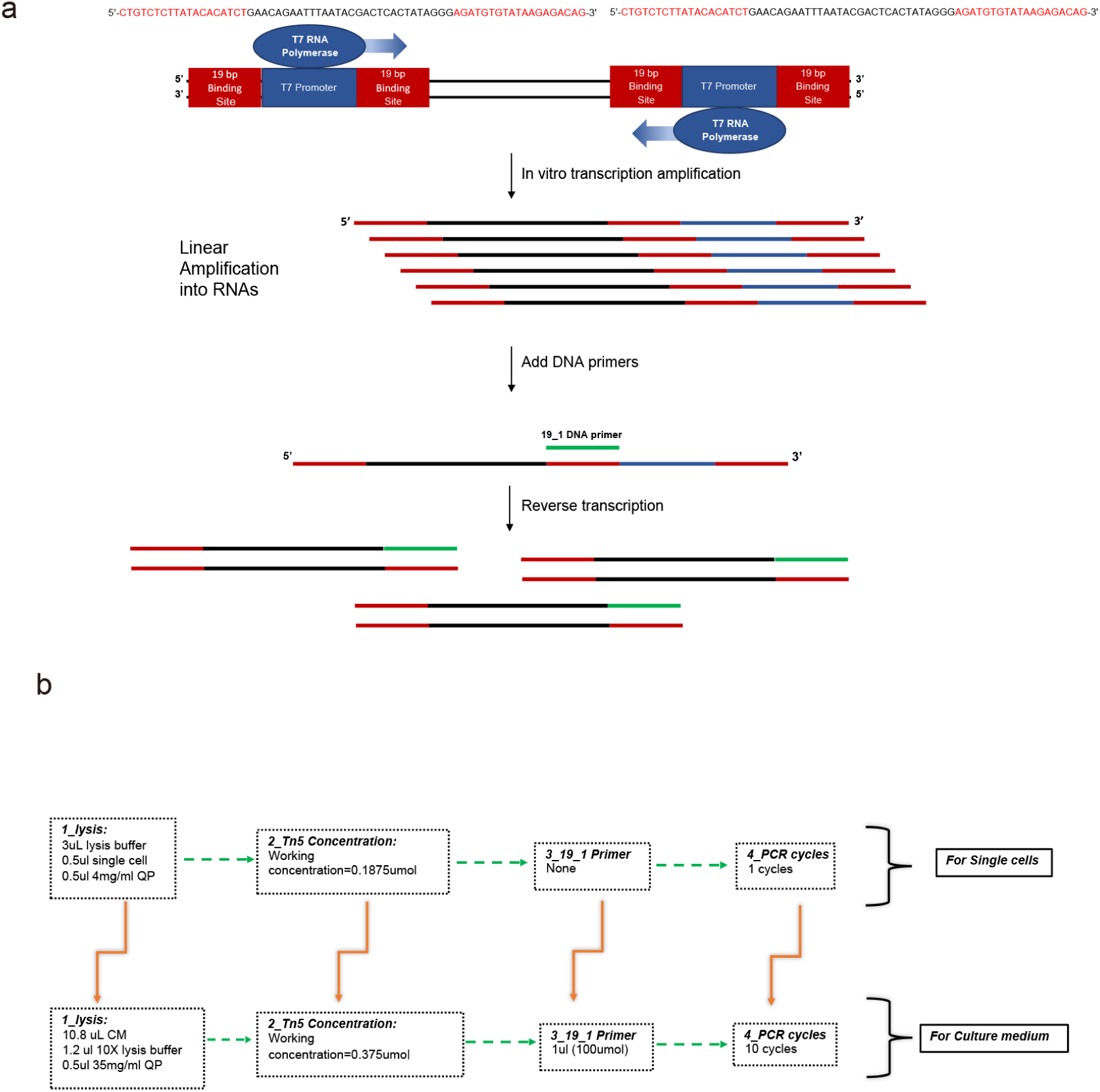
Improved LIANTI protocol. In this study, we used a modified version of the LIANTI single-cell amplification method, which involves 1) Transposon-assisted DNA fragmentation; 2) Gap filling and extension; 3) Reverse transcription; 4) Second-strand synthesis; 5) Library construction and sequencing. To adapt this method for culture medium samples, we made the following improvements: 1) Adjusted the lysis system to a 10.8 µl sample volume and increased QIAGEN Protease (Qp) concentration to 35 mg/ml; 2) Doubled the transposon concentration; 3) Added a 19-bp transposon ME sequence complementary primer to enhance reverse transcription efficiency; 4) Included 10 additional PCR cycles in second-strand synthesis to improve the conversion of single-stranded cDNA to double-stranded DNA.

**Extended Data Fig. 3.**
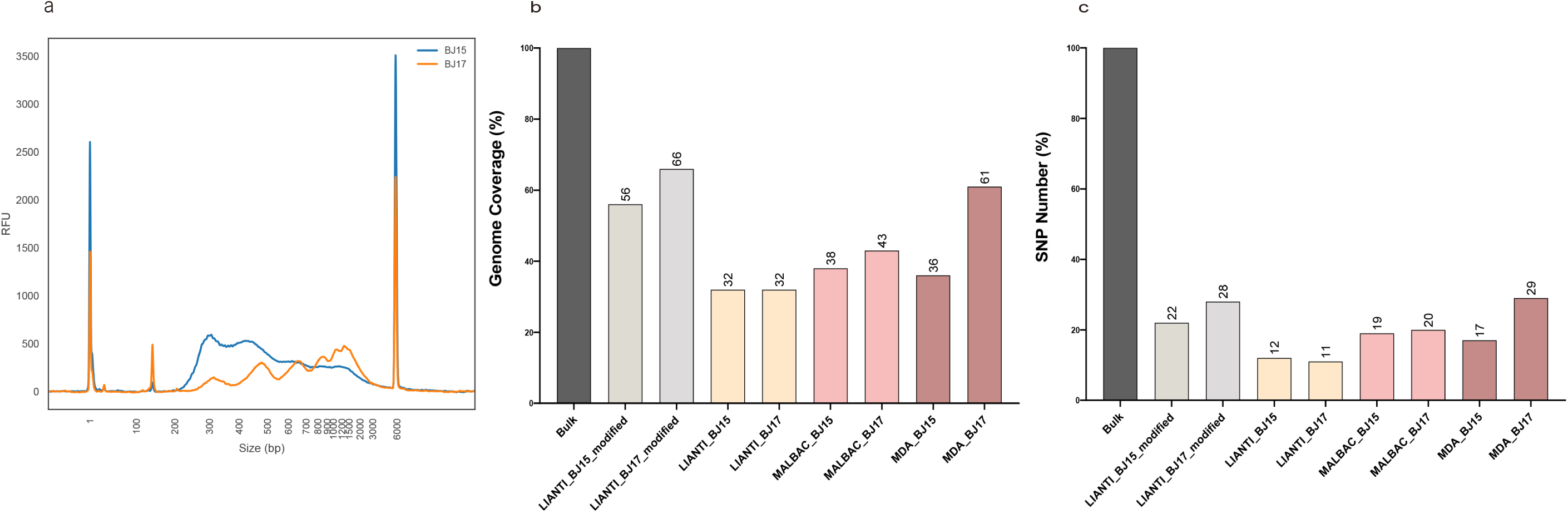
Improved LIANTI amplification for culture medium samples. For method development we utilized the BJ cell line to simulate the embryo culture process. BJ15 medium was collected on day 5, BJ17 on day 7. a) Size distributions of DNA molecules in BJ15 and BJ17 cell culture medium (Agilent Fragment Analyzer System). b) Genome coverage analysis of BJ15 and BJ17 cell culture medium, amplified using Improved LIANTI, MALBAC, and MDA. c) SNP number analysis of BJ15 and BJ17 cell culture medium, amplified using Improved LIANTI, MALBAC, and MDA. Genome coverage and SNP number normalization calculations were carried out relative to the Bulk sample. The sequencing depth for bulk genomic DNA (gDNA) exceeds 30X, while for amplified products, the sequencing depth exceeds 10X. See Supplementary methods and Table S2 for more details.

**Extended Data Fig. 4.**
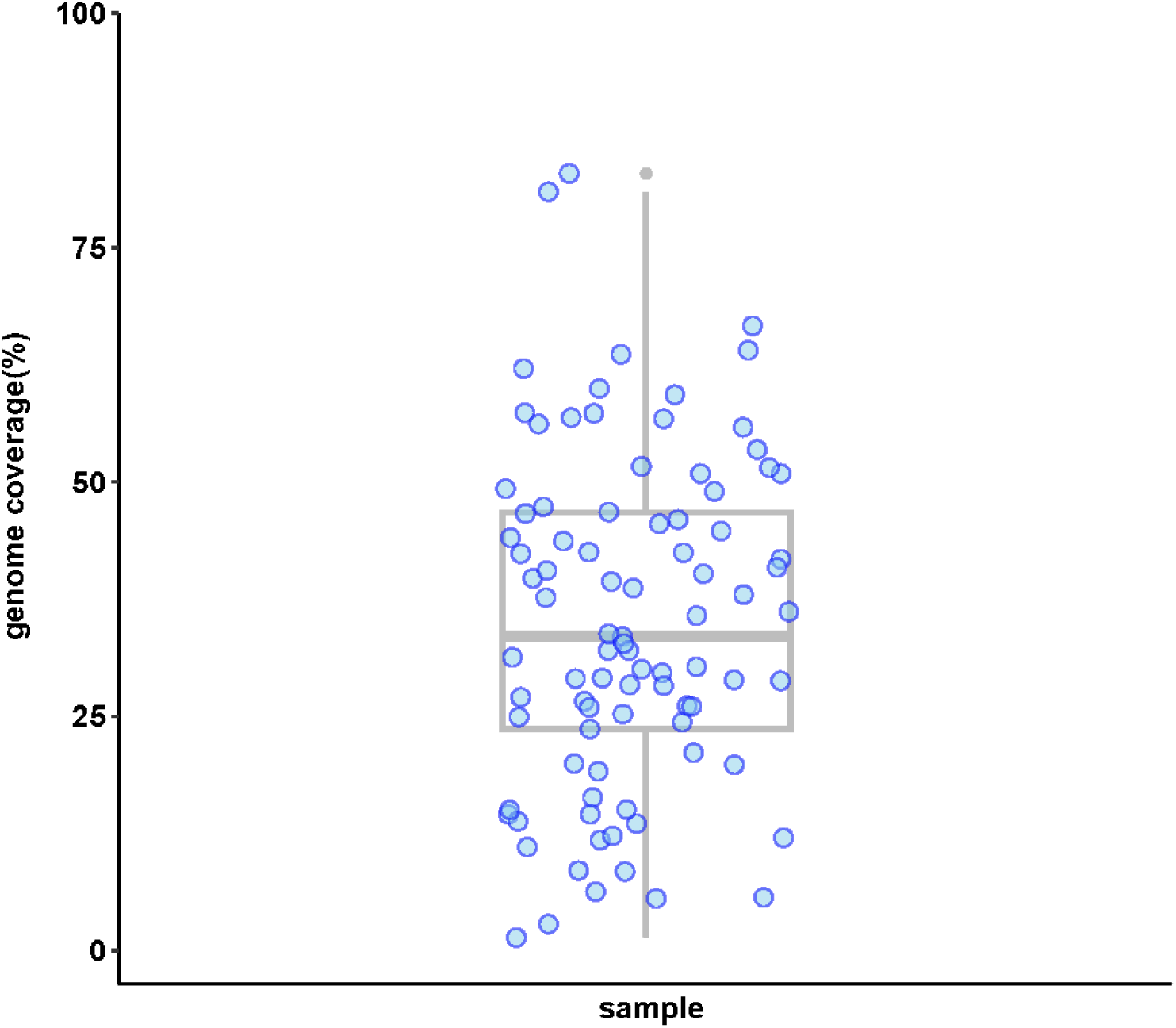
Genome coverage of spent embryo culture medium samples. Genome coverage of all SCM samples from Clinical Center A. Data points are spread along the x-axis for visualization only; position along the x-axis does not carry meaning. Thick gray line is Median, with quartiles represented by the box and the whiskers.

**Extended Data Fig. 5.**
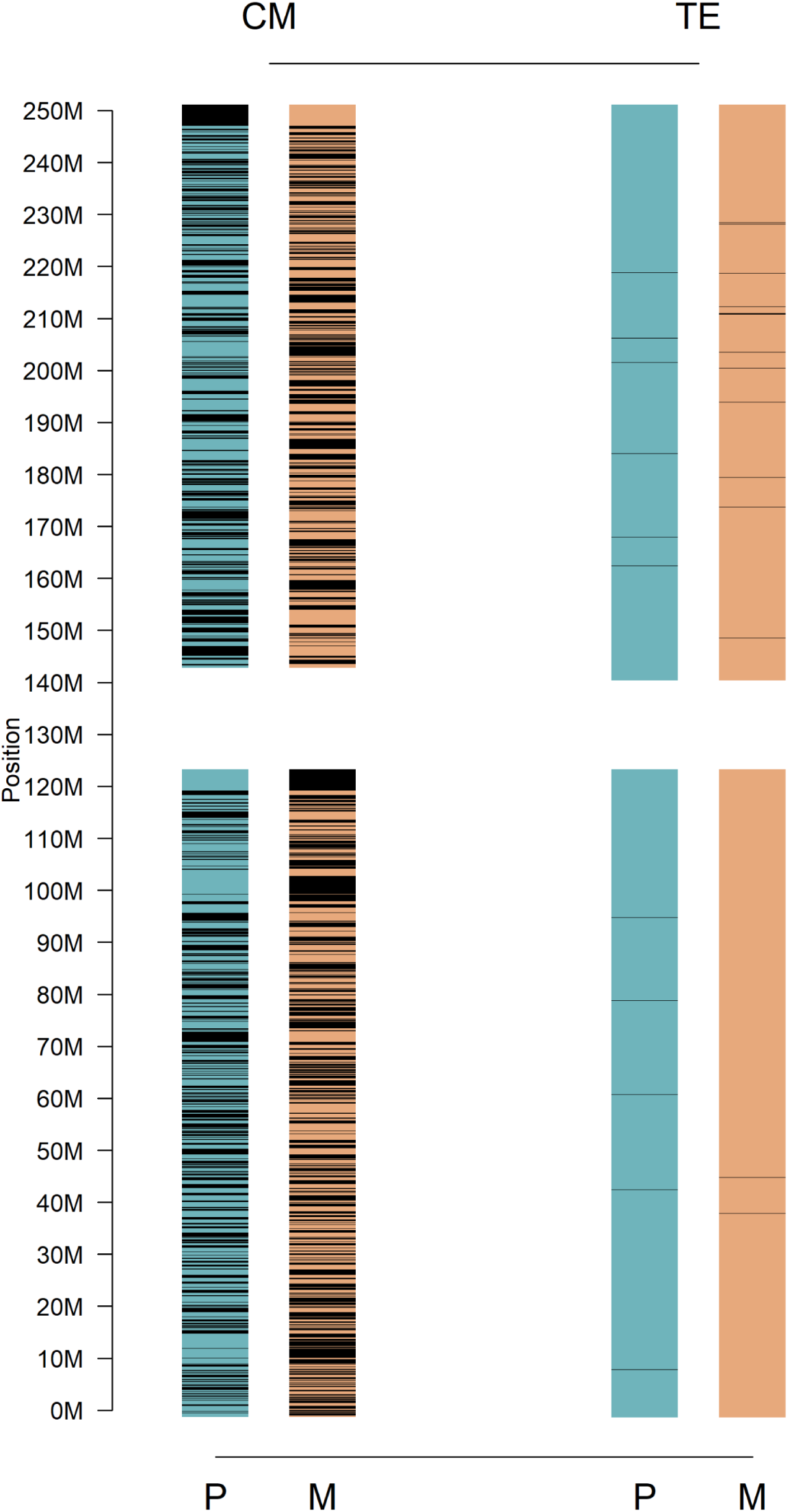
Illustration of large fragment loss of haplotype from Chromosome 1 of a single spent embryo culture medium sample. CM culture medium; TE Trophectoderm biopsy. Blue and orange bars represent haplotypes from paternal and maternal chromosome 1 respectively. Black regions represent loci with no genetic information observed in specific haplotypes.

**Extended Data Fig. 6.**
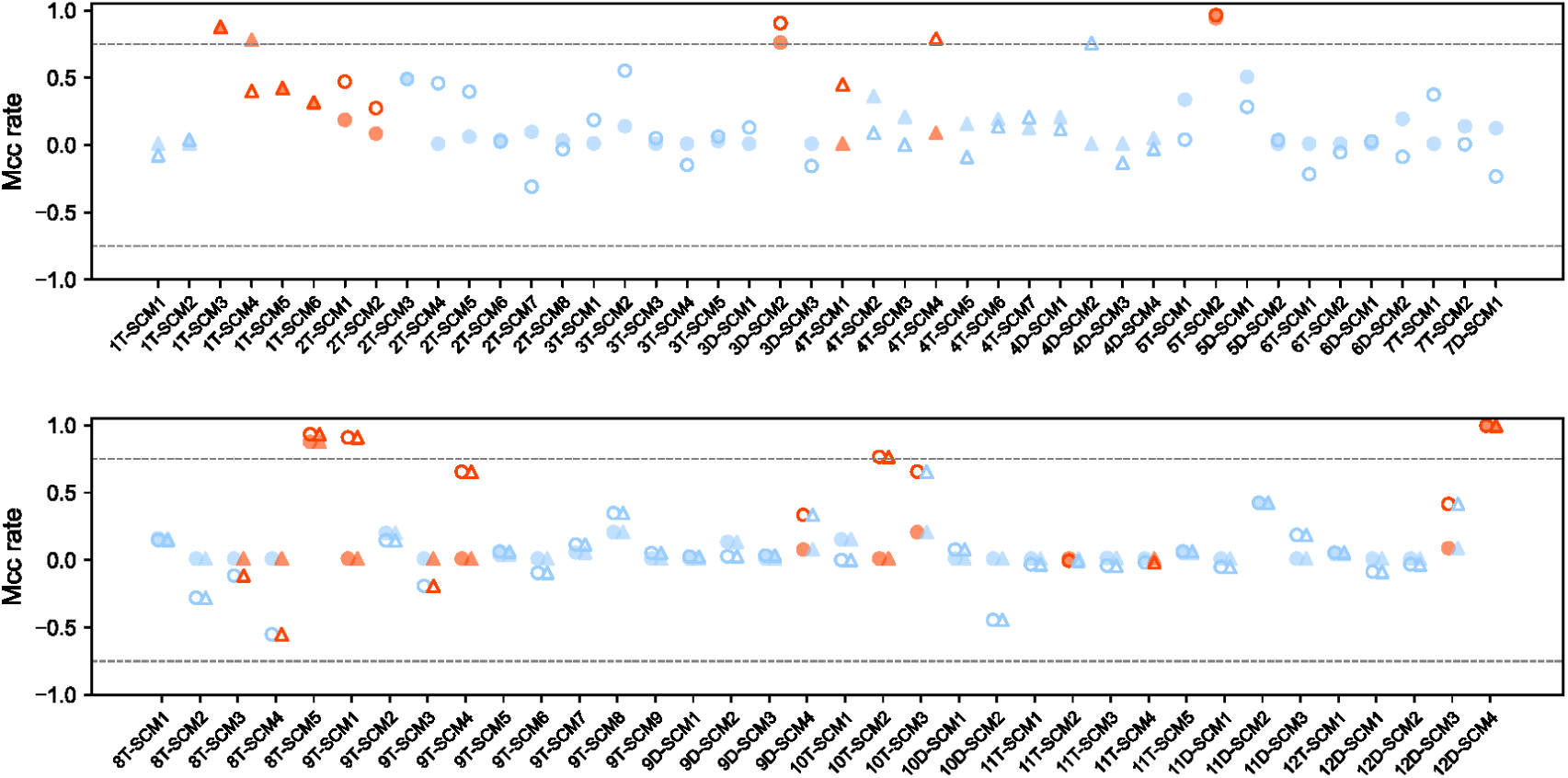
Estimated maternal cell contamination rates. Maternal cell contamination rates for autosomal dominant (top panel) and autosomal recessive family samples (bottom panel) from Clinical Center A. Circles represent pathogenic loci inherited paternally, while triangles denote maternal inheritance. Open symbols indicate the initially estimated rates, and solid symbols represent the final rates after performing the iterative algorithm in our Bayesian linkage analysis method. Blue symbols correspond to diagnosable samples, whereas red symbols denote undiagnosable samples. See Table S3 for the source data.

**Extended Data Fig. 7.**
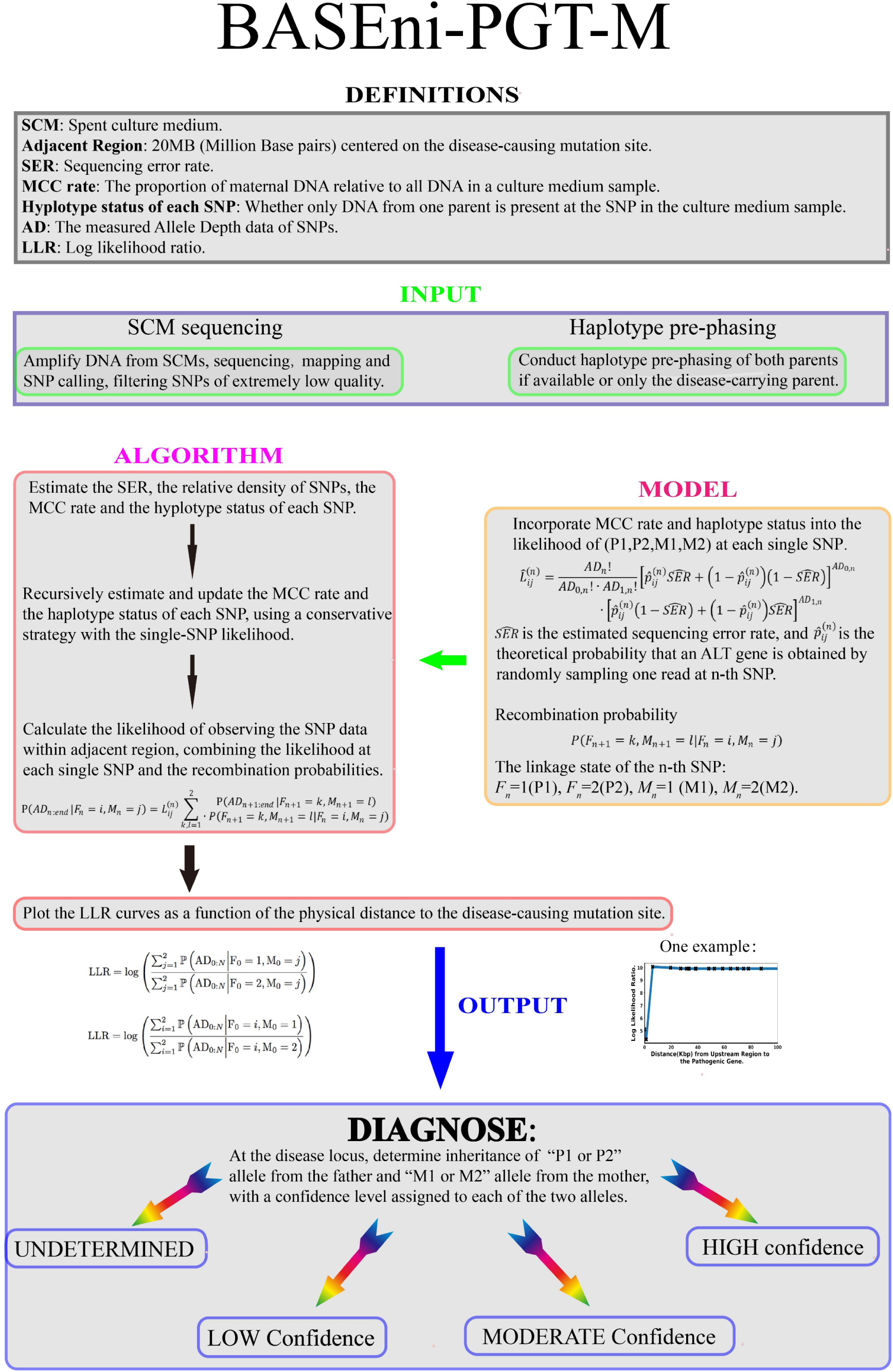
Bayesian linkage analysis workflow. The Bayesian linkage-analysis model uses sequencing data from SCM and the phased haplotypes of the parents as input. By iteratively estimating the maternal cell contamination rate and haplotype states of each SNP, the model calculates the log-likelihood ratio of inheriting disease-carrying alleles versus disease-free alleles as a function of distance to the disease-causing mutation site. This curve, with its distinct characteristics, allows classification of the diagnosis into four confidence categories: High, Moderate, Low, and Undetermined.

**Extended Data Fig. 8.**
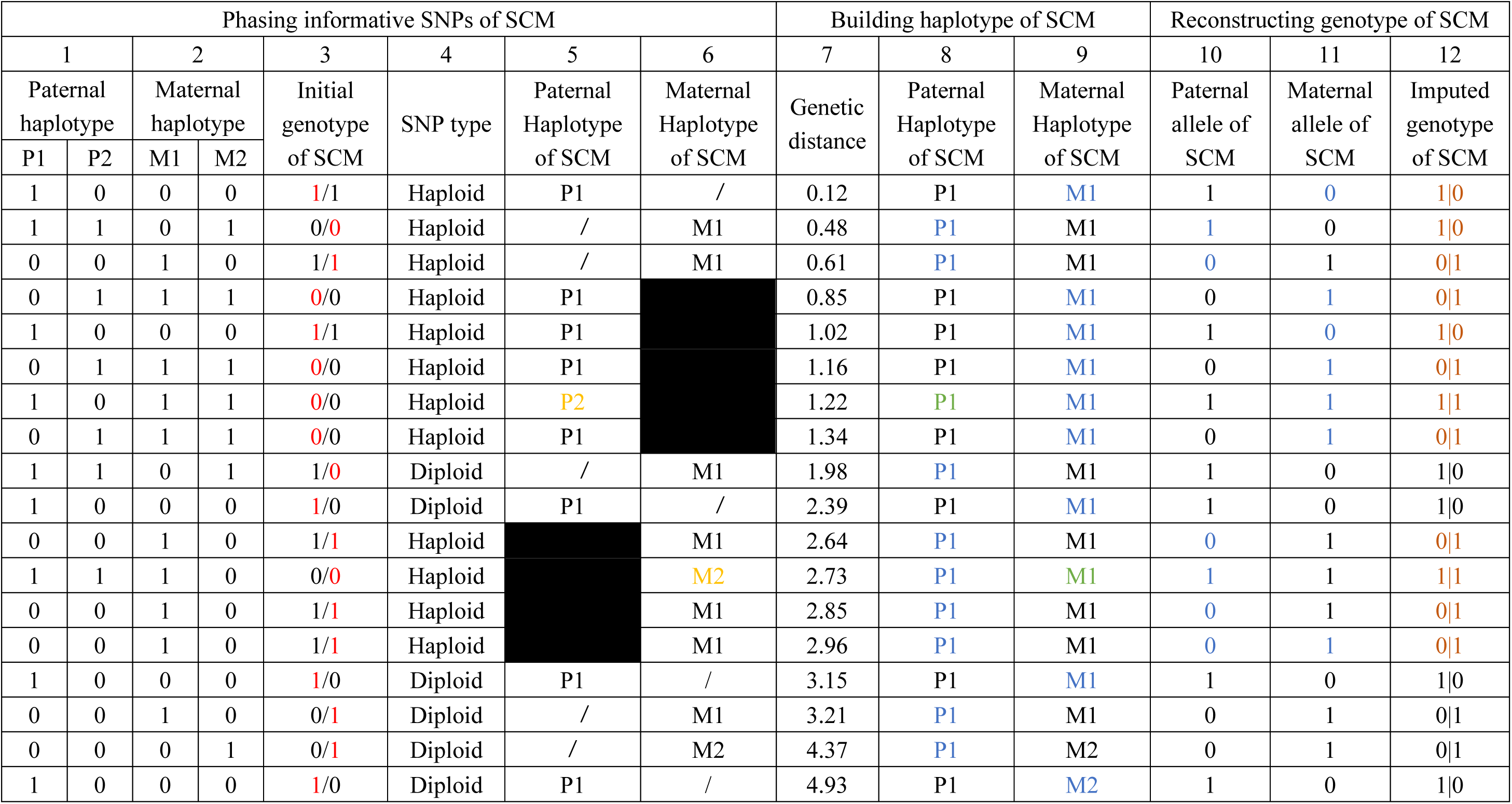
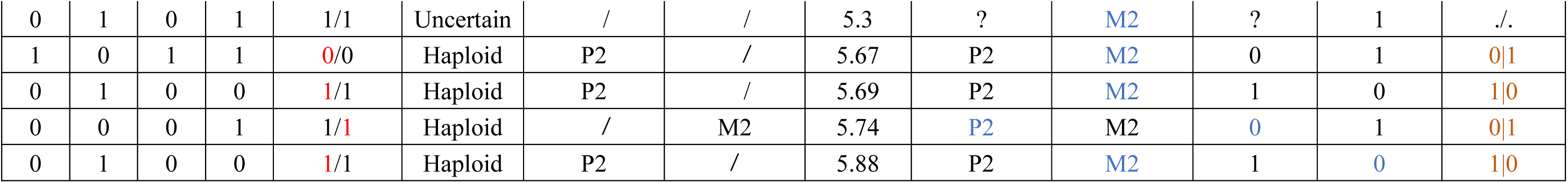
An example workflow of pedigree-based whole genome reconstruction. The format of all genotypes is (P/M). In the initial genotype of the spent embryo culture medium (column 3; see second row for column number), red numbers indicate informative alleles, defined as alleles which could only have originated from one of the parents based on the phased parental haplotypes (columns 1-2). Slashes in columns 5 and 6 (“Paternal Haplotype of SCM” and “Maternal Haplotype of SCM”) indicate haplotypes that were undetermined in the initial reconstruction. Black filled cells represent a large haplotype fragment loss, characterized by consecutive SNPs missing the same haplotype. Yellow text in these columns indicate incorrectly inferred SCM haplotypes, which were corrected by the Hidden Markov Model method, as shown by green text in columns 8 and 9. In columns 8-11, blue text denotes imputed haplotypes on non-informative alleles. Brown numbers in column 12 show the final imputed allele or genotype of the genome.

**Extended Data Fig. 9.**
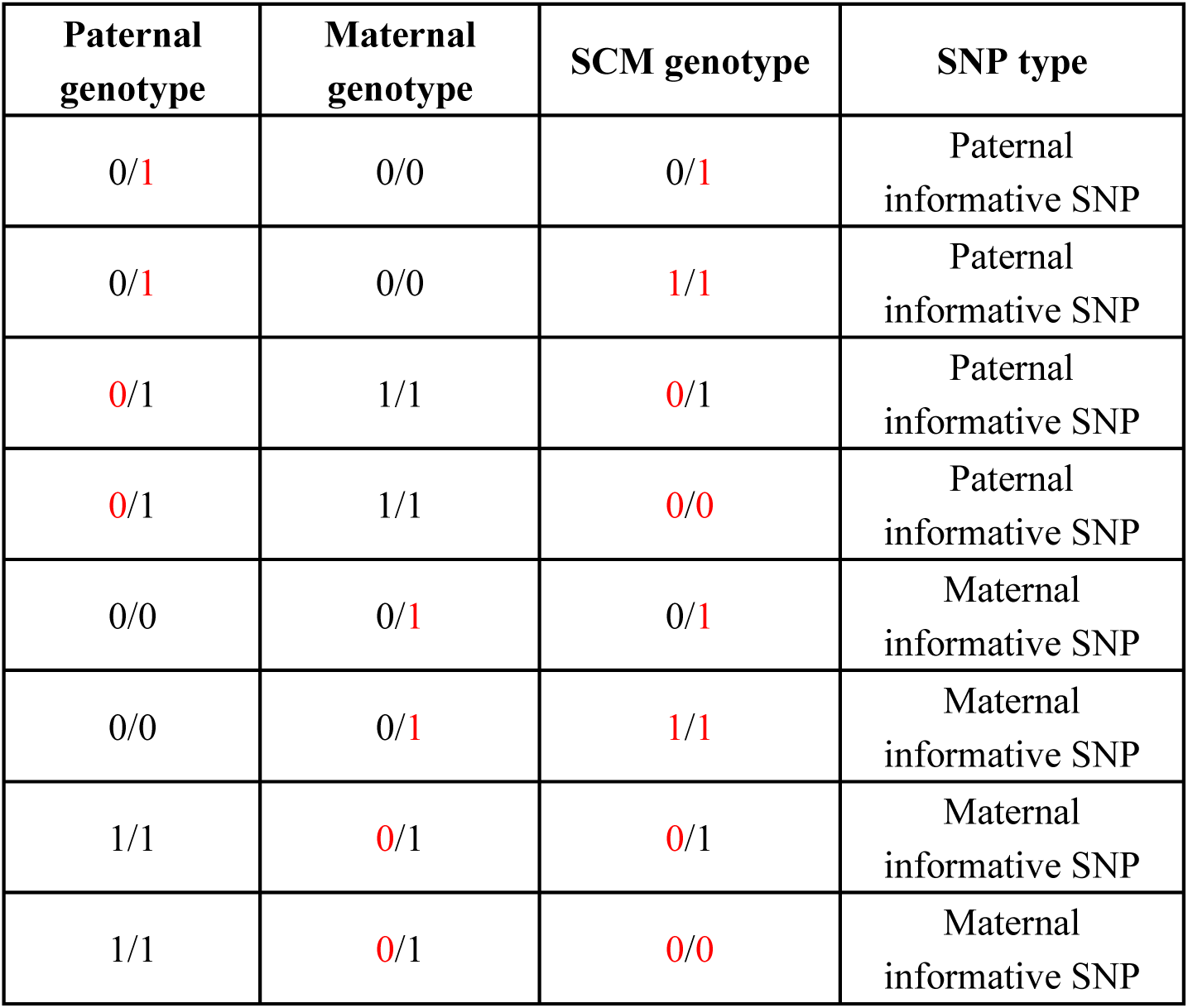
Strategy for informative SNPs selection. 0 and 1 respectively denote reference and alternative alleles of variant. In the first two columns, red text indicates informative SNPs, defined as alleles which could only have originated from one of the parents based on the phased parental haplotypes. These alleles are also denoted by red text in the initial SCM genotype determination (column 3), although that determination is made independent of information about the parental haplotypes and is therefore uncorrected in this chart, resulting in some impossible genotypes.

