## Supplementary methods for "Highly Accurate Non-Invasive Preimplantation Genetic Testing for Monogenic and Polygenic Diseases from Spent Medium"

1. Experimental methods

2. Linkage analysis methods

3. Whole genome construction methods

### 1 Supplementary Experimental Methods

**DNA Fragment Length Distribution in Spent Culture Medium**

A major limitation encountered in studying DNA fragment length was the trace amounts of genetic material available. DNA amplification followed by library preparation emerged as the primary approach. However, potential biases in DNA amplification, attributable to DNA size, might impact the accuracy of size determination and subsequent downstream amplifications. In this study, we opted for the Accel-NGS® 2S DNA Library Kit (Swift Biosciences, USA) to directly construct sequencing libraries from 2μL of spent blastocyst medium. We strictly adhered to the detailed operational steps provided by the manufacturer. Briefly, the process involved Repair I, Post-Repair I purification, Repair II, Post-Repair II purification, Ligation I, Post-Ligation I purification, Ligation II, Post-Ligation II purification, and PCR enrichment. The thermal cycling conditions for PCR enrichment were as follows: 45s at 98℃, followed by 15 cycles at 98℃ for 15s, 60℃ for 30s, and 72℃ for 60s, with a final step at 72℃ for 60s and then held at 4℃. Six spent blastocyst media were utilized to investigate the DNA size distribution in spent embryo culture medium samples. Subsequently, the distribution of library insert fragments was analyzed in six constructed libraries using the Agilent 5200 fragment analyzer.

**MDA, MALBAC, and improved LIANTI Amplification of Two Simulated Embryo Culture Medium (Shorter and longer DNA Fragments)**

In this study, we used the BJ primary human foreskin fibroblast (CRL-2522™, ATCC) cell line, a diploid human cell line, to culture and obtain cell-free DNA from the culture medium at different time points. The culture medium of BJ cell line was used as a standard sample for comparison of different single-cell whole genome amplification methods [1]. This was done to simulate the size distribution of DNA fragments in blastocyst culture medium at various culture stages (Accel-NGS® 2S DNA Library Kit, Swift Biosciences, USA). After thawing the BJ cells stored in liquid nitrogen, they were cultured in Eagle's Minimum Essential Medium (EMEM) supplemented with 10% Fetal Bovine Serum (FBS, Gibco) and 1% Penicillin-Streptomycin (PS, Gibco). The cell culture conditions were as follows: humidity = 95%, temperature = 37°C, and CO2 concentration = 5%. Both the BJ cells and EMEM (30-2003™, ATCC) were purchased from ATCC (https://www.atcc.org/). Fetal Bovine Serum, qualified, Australia (10099141, Gibco), and Penicillin-Streptomycin (15140122, Gibco) were purchased from Gibco (https://www.thermofisher.cn/).

The BJ cells, which are adherent, were subjected to a medium change on the third day after revival. The procedure was as follows: the cell culture medium was collected into a 15ml centrifuge tube, and a fresh culture medium was added. The collected medium was centrifuged at 1800 rpm for 5 minutes, and the top 2/3 of the supernatant was transferred to a 1.5ml centrifuge tube and stored at -20°C. On the fifth day after revival, the cell culture medium was collected into a 15ml centrifuge tube. The collected medium on the fifth day was processed in the same way as the medium collected on the third day.

MDA (Qiagen, REPLI-g Advanced DNA Single Cell Kit), MALBAC (Yikon Genomics, China), and Improved LIANTI (developed in this paper) were employed to amplify the collected culture medium. Deep sequencing was used to assess the amplification efficiency of different methods on simulated samples.

**Amplification Bias Evaluation Caused by DNA Size Difference**

Three whole genome amplification methods, including MDA, MALBAC, and Improved LIANTI were included in this study. We designed two lengths of PCR products (186bp primer, Forward: 5’-TGGAGGTAGTAGAGCCTGAAGTC-3’; Reverse: 5’-TCCTTATCACCTTCATAGAAAG-3’ and 1305bp primer, Forward: 5’-TGACTGCAGAGGCAAGTGAT-3’; Reverse: 5’-

AGAACAGCAGGAGGGATAAACTAACC-3’), which were mixed together in equimolar proportions. After the dilution of the mixture, 40 pg of the template was taken, and amplification was performed using MDA (Qiagen, REPLI-g Advanced DNA Single Cell Kit), MALBAC (Yikon Genomics, China), and Improved LIANTI (developed in our lab). Following the fragmentation of the products and bulk samples, library construction was conducted using the NEBNext® Ultra™ II DNA Library Prep Kit for Illumina (NEB, USA). Statistical analysis was performed to determine the number of reads from the sequencing data originating from the two regions.

### 2. Linkage analysis method

Previous linkage analysis methods are based on the Lander-Green algorithm and its variants, which do not consider maternal cell contamination and require high quality sequencing data [2-6]. Directly applying these methods results in high rates of misdiagnosis [7,8]. There are a few linkage analysis methods that have taken into account the mixing or contamination with maternal cells [9,10]. However, these methods still require much higher quality sequencing data than those from culture media, and their main applications are based on blood samples or cells in the placenta or amniotic fluid of the mother during pregnancy [9,10]. These existing methods will cause high diagnostic errors when applied to the sequencing data from spent embryo culture media.

We provide a linkage analysis method called BASE-niPGT (BAyesian linkage analySis mEthod for non-invasive Preimplantation Genetic Testing of Monogenic disorders) that addresses the problems of large fragment haplotype loss, low coverage, and high fraction of maternal cell contamination (MCC) that may hinder the inference of inherited parental haplotypes at the disease-causing mutation site. The method contains the following main steps: a) Calculating the likelihood at each single SNP incorporating the MCC rate and haplotype status of the SNP; (b) Recursively estimating the MCC rate and the haplotype status of each SNP; (c) Calculating the likelihood of observing the SNP data within regions adjacent to the disease- causing mutation site; (d) Determining whether the embryo carries the disease-causing haplotype and providing a level of confidence.

The Bayesian model takes the sequencing data, as well as the phased parents' haplotypes, as input. To enhance the data quality, SNPs of extremely low quality are filtered out. Several parameters were estimated before calculating the likelihood ratio: the sequencing error rate, MCC rate and haplotype status. MCC rate was defined as the fraction of DNA fragments sourcing from the maternal chromosome. The haplotype status was defined as the true parental source of the DNA at each SNP and was divided into 'Paternal Chromosome Only', 'Maternal Chromosome Only', or 'Both Parental Chromosomes'. Parental Chromosome Only, for instance, suggested that the DNA detected at this SNP solely came from the father. MCC rate and haplotype status were repeatedly calibrated until convergence.

Then, the likelihood of sequencing data within a specific physical distance to the disease-causing mutation site was calculated. The recombination probability can be obtained from a long-existing dataset such as [11], while the single-SNP likelihood was attained from a binomial model, whose parameters were determined by the estimated sequencing error rate, MCC rate and haplotype status.

Instead of considering a fixed number of SNPs, as has been the case in previous studies, our method adds SNPs one by one starting from the disease-causing mutation site and calculates the log-likelihood ratio for each SNP subset, which results in a curve of the log-likelihood ratio against the physical distance of each added SNP. Typically, the curve starts at a point where the ordinate is close to zero and eventually stabilizes at a plateau as a sufficient number of SNPs are added. Based on the characteristics of the curve one can determine which haplotype had been inherited, along with a confidence level, which is classified into four categories: High Confidence, Moderate Confidence, Low Confidence and Undetermined. Using this method, alleles can be accurately distinguished even in the presence of maternal contamination.

**Detailed description of BASE-niPGT**

Step 1): Amplifying DNA molecules from the *in vitro* embryo culture medium. Preparing DNA libraries and sequencing the DNA molecules. Performing mapping and SNP calling to generate the SNP data and controlling its quality.

All SNPs with DP values less than 5, or QUAL less than 30, or genotype quality (GQ) values less than 10 in the blood samples are excluded, because they are considered to be of too poor quality and too small signal-to-noise ratio. For the cultural medium sample, we excluded the 20% SNP with lowest GQ values as well as all the SNP with GQ<=3. The SCM samples with low GQ(<=3) rate larger than 0.4 will also be ruled out.

The "adjacent region" is defined as the 10MB (Million Base pairs) upstream and downstream of the disease-causing mutation site. After removing low-quality SNPs, the Sequencing Error Rate (SER) in the adjacent region is estimated. SNPs with genotype 0|0 of both father and mother, or genotype 1|1 of both father and mother are selected. Ideally, at such SNPs, genotypes that are identical to the parents' genotype should only be detected. Therefore, sequencing data with genotypes different from those of the parents must be errors. Hence, the estimated SER is defined as the proportion of the total number of erroneous reads versus the total number of reads.

The relative density of SNPs detected in the adjacent region of the disease-causing mutation site is estimated, and defined as the ratio of the total number of SNPs detected in this area compared to the total number of human common SNPs within this region. Here, human common SNPs are defined as all SNPs with Minor Allele Frequency (MAF) greater than or equal to 0.01. The Minor Allele Frequency of a group of alleles refers to the frequency of its second-most frequent allele in a population. The human common SNP data updated in 2018 by dbSNP [12] was used to calculate the relative SNP density for each sample.

After completing the above steps, samples of culture medium with low SNP density (<0.001) or a high sequencing error rate (>0.2) are considered to be of extremely low quality and thus excluded. Such samples are categorized as "Rule out" due to low data quality.

Step 2): Conducting haplotype pre-phasing, for the blood samples of both parents or solely the disease-carrying parent.

To deduce parental haplotypes, genomic data from the father and mother of the embryo and from any living siblings of the embryo, and alternatively or in addition, from parental siblings, grandparents, or discarded embryos. Discarded embryos samples require amplification, which may introduce errors. Data from several qualified discarded embryo data may help to increase the accuracy of pre-phasing.

Plink 1.9 with the option "--mendel" is used to identify sites with Mendelian errors. Additionally, merlin [2] or BEAGLE 4.0 [13] is used to phase the genotype data, taking into account the parents-offspring relationships and simultaneously eliminating sites that contain Mendelian errors.

<u>Step 3): Incorporating the MCC rate and haplotype status into the likelihood of the four possible inheritance scenarios (P1, P2, M1, or M2) at each single SNP.</u>

For single-SNP likelihood, the binomial distribution is used to build a model, where the parameters of the binomial distribution used were the estimated sequencing error rate $\widehat{SER}$, MCC rate $\hat{\alpha}$, and detected allele depth (AD) values of the two alleles at each single SNP.

Specifically, the following

$$l_{11} = \left(\frac{1-\hat{\alpha}}{2}, 0, \frac{1}{2}, \frac{\hat{\alpha}}{2}\right)^T, \quad l_{12} = \left(\frac{1-\hat{\alpha}}{2}, 0, \frac{\hat{\alpha}}{2}, \frac{1}{2}\right)^T$$

$$l_{21} = \left(0, \frac{1-\hat{\alpha}}{2}, \frac{1}{2}, \frac{\hat{\alpha}}{2}\right)^T, \quad l_{22} = \left(0, \frac{1-\hat{\alpha}}{2}, \frac{\hat{\alpha}}{2}, \frac{1}{2}\right)^T$$

defines as the theoretical fraction of four alleles ($P_1$, $P_2$, $M_1$, $M_2$) in embryonic culture medium. To take the status of haplotype into consideration, define

$$h(H) = \begin{cases} (1,1,0,0)^T & if\ H = P \\ (0,0,1,1)^T & if\ H = M \\ (1,1,1,1)^T & if\ H = PM\ or\ H = Unknown \\ (1,0,0,0)^T & if\ H = P_1 \\ (0,1,0,0)^T & if\ H = P_2 \end{cases}$$

where H is the haplotype status at this SNP (See step 6 for more information about haplotype status).

Then for the n-th SNP we considered, define

$$\hat{l}_{ij} = \frac{l_{ij} \circ h(H_n)}{l_{ij}^T h(H_n)}$$

Here ◦ denotes product by position and $H_n$ is the haplotype status at the n-th SNP. Paternal genotype at the n-th SNP is denoted as pGT = $pGT_1/pGT_2$ and the maternal one as mGT = $mGT_1/mGT_2$, where $pGT_1$, $pGT_2$, $mGT_1$, $mGT_2$ $\in \{0,1\}$.

Then define $GT_n = (pGT_1, pGT_2, mGT_1, mGT_2)$, and denotes $\hat{p}_{ij}^{(n)} = \hat{l}_{ij}^T \cdot GT_n$, which is the theoretical probability that an ALT gene is obtained by randomly sampling one read at n-th SNP in the culture medium when MCC rate and haplotype loss are taken into consideration. Therefore, the single-SNP likelihood is formulated as

$$\hat{L}_{ij}^{(n)} = \frac{AD_n!}{AD_{0,n}! \cdot AD_{1,n}!} \left[\hat{p}_{ij}^{(n)} \widehat{SER} + \left(1 - \hat{p}_{ij}^{(n)}\right)\left(1 - \widehat{SER}\right)\right]^{AD_{0,n}} \cdot \left[\hat{p}_{ij}^{(n)}\left(1 - \widehat{SER}\right) + \left(1 - \hat{p}_{ij}^{(n)}\right)\widehat{SER}\right]^{AD_{1,n}}$$

for i, j=1, 2.

Step 4): Estimating the MCC rate, and determining the haplotype status of each SNP to represent whether only DNA from one parent is present in the culture medium.

Step (4a): Initially estimating the MCC rate and haplotype status of each SNP in the region adjacent to the disease-causing mutation site.

The MCC rate is defined as the proportion of maternal DNA in the culture medium of an embryo compared to the total amount of DNA and this value should theoretically be between 0 and 1. All SNPs with paternal genotype 0|0 and maternal genotype 1|1, or with paternal genotype 1|1 and maternal genotype 0|0 in the adjacent region are selected. At these SNPs, it is determined theoretically whether each read detected in the culture medium is from the father or the mother, and an initial estimate of MCC rate can be obtained by subtracting the paternal reads from the maternal reads and then dividing by the total number of reads. If the MCC rate is negative, we set it to 0.01, as such negative values are likely attributable to sequencing error or loss of the maternal allele.

The haplotype status of SNPs that can be directly determined from their AD (Allele Depth) values are first labeled as either containing only paternal DNA (P), only maternal DNA (M), or both (PM). Others remain unlabeled.

Then, the surrounding SNPs in the upstream and downstream of these labeled SNPs are searched, and it is then determined whether the surrounding SNPs could be labeled with the same label as the labeled SNP in the first step.

The parameter T used below is assumed to be an integer threshold determined in advance (the default value is 3).

For SNP in the region adjacent to the disease-causing mutation site, those satisfying one of the following conditions are labeled as P.

• The paternal genotype is 0|0 and the maternal genotype is 1|1. $AD_0 > T$ and $AD_1 = 0$.

• The paternal genotype is 1|1 and the maternal genotype is 0|0. $AD_1 > T$ and $AD_0 = 0$.

• The paternal genotype is 0|1 and the maternal genotype is 0|0. $AD_0 = 0$ and $AD_1 > T$.

• The paternal genotype is 0|1 and the maternal genotype is 1|1. $AD_0 > T$ and $AD_1 = 0$.

Some SNPs labeled as P can be further labeled as $P_1$ or $P_2$, indicating that the detected SNP is from the paternal source allele or the maternal source allele of the father. Specifically, SNP labeled P is labeled as $P_1$, if the SNP satisfies one of the following.

• The paternal genotype is 1|0 and the maternal genotype is 0|0. $AD_1 > T$ and $AD_0 = 0$.

• The paternal genotype is 0|1 and the maternal genotype is 1|1. $AD_0 > T$ and $AD_1 = 0$,

where 0|1 and 1|0 denote the paternal genotype, and the number at left-hand-side and right-hand-side of | denote the paternal source and maternal source gene of the father at this SNP respectively.

Similarly, a SNP labeled P is labeled as $P_2$, if the SNP satisfies one of

the following two conditions.
• The paternal genotype is 0|1 and the maternal genotype is 0|0. $AD_1 > T$ and $AD_0 = 0$.
• The paternal genotype is 1|0 and the maternal genotype is 1|1. $AD_0 > T$ and $AD_1 = 0$,

SNPs satisfying one of the following conditions are labeled as M.
• The paternal genotype is 0|0 and the maternal genotype is 1|1. $AD_1 > T$ and $AD_0 = 0$.
• The paternal genotype is 1|1 and the maternal genotype is 0|0. $AD_0 > T$ and $AD_1 = 0$.

Due to the existence of MCC, no SNP is labeled as M1 or M2. SNPs satisfying one of the following conditions are labeled as PM.
• The paternal genotype is 0|0 and the maternal genotype is 1|1. $AD_1 > [T/2]$ and $AD_0 > [T/2]$;
• The paternal genotype is 0|0 and the maternal genotype is 1|1. $AD_1 > [T/2]$ and $AD_0 > [T/2]$;
• The paternal genotype is 0|1 and the maternal genotype is 1|1. $AD_1 > [T/2]$ and $AD_0 > [T/2]$;
• The paternal genotype is 0|1 and the maternal genotype is 0|0. $AD_1 > [T/2]$ and $AD_0 > [T/2]$;
where [] denotes the floor function.

After completing the initial labeling procedure, there are 5 possible labels: P, M, PM, $P_1$, $P_2$. Suppose there are several labeled SNPs among the adjacent region of the disease-causing mutation site, with an ordinal number $-N_1 \leq s_1 < s_2 < \dots < s_D \leq N_2$. Then, for a particular SNP $s_d$ that has been labeled, a search is carried out between the SNPs with ordinal number $s_d-1$ and $s_d+1$. The $s_d$-th SNP is called the central SNP and the region between $s_d-1$-th and $s_d+1$-th SNP is called the searching region. The search region is divided into two parts: the upstream part and the downstream part of the $s_d$-th SNP. In the downstream searching, each SNP in the downstream searching region is assigned a label. The n-th SNP during downstream searching from its nearest labeled SNP upstream from it is denoted as $H_n^{down}$ and during upstream searching from its nearest labeled SNP downstream from it is denoted as $H_n^{up}$ .

The search process only in the downstream region is described as follows as an example. First, $H_{s_d}^{down}$ is initialized as the label assigned in the first labeling step. Next, assume $s_d$, $s_d$ +1, ..., $s_d$ +m−1 are all labeled the same as $H_{s_d}^{down}$. The next step is to search the ($s_d$ + m)-th SNP and determine whether the same label can be assigned to this SNP. The same label is assigned to ($s_d$+m)-th SNP if and only if

$$\frac{\sum_{i,j=1}^{2} \mathbb{P}\left(AD_{s_d:s_d+m} | P_{s_d} = i, M_{s_d} = j, H_{s_d:s_d+m}^{down} = H_{s_d}^{down}\right)}{\sum_H \sum_{i,j=1}^{2} \mathbb{P}\left(AD_{s_d:s_d+m} | P_{s_d} = i, M_{s_d} = j, H_{s_d:s_d+m-1}^{down} = H_{s_d+m}^{down} = H\right)} \geq \lambda_0$$

Here, $H_{s_d:s_d+m}^{down} = H_{s_d}^{down}$ denotes $H_l^{down} = H_{s_d}^{down}$ , for any $l = s_d, s_d + 1, \cdots, s_d + m$, and $\lambda_0$ is a pre-determined threshold. H in the summation refers to summing over H = P, M, PM, $P_1$, $P_2$ and $AD_{s_d:s_d+m}$ denotes $\{AD_{s_d}$ , $AD_{s_{d+1}}$, ..., $AD_{s_d+m}\}$. $P(\cdot \mid \cdot)$refers to the conditional probability, specifically, the probability of the observed AD values at the $s_d$-th SNP, conditioned on the knowledge of the inherited haplotype from both parents at the $s_d$-th SNP as well as the labels of nearby SNPs. The conditional probability is calculated in exactly the same way as the recursion elaborated on in the next section. For each central SNP, the haplotype state for at most 10 SNPs upstream or downstream is inferred.

As a result of the searching procedure, there will be at most two labels at each SNP. For those SNPs with less than two labels, a label is added as "Unknown". Then, the two labels of each SNP are merged to finalize the labeling procedure of haplotype status. According to the principle of final labeling, a SNP is labeled as P or M only after sufficient confidence that only paternal or maternal DNA fragments are detected, otherwise the SNP is labeled as PM. Specifically, the final label of the n-th SNP is

$$H_n = \begin{cases} P & if\ \{H_n^{up}, H_n^{down}\} = \{P_1\}\ or\ \{P_1, P\}\ or\ \{P, PM\}\ or\ \{P_1, Unknown\} \\ & or\ \{P_2, P\}\ or\ \{P_2, PM\}\ or\ \{P_2, Unknown\} \\ M & if\ \{H_n^{up}, H_n^{down}\} = \{M\}\ or\ \{M, Unknown\} \\ PM & otherwise \end{cases}$$

Step (4b): Recursively updating the MCC rate and haplotype status of each SNP

Analyzing the haplotype status of each SNP can provide information regarding whether parental DNA fragments are detected at each SNP, which can facilitate a re-estimation of MCC rate. With the new estimate of MCC rate, the likelihood will change and the final labels at all SNPs should be updated. Since the labels at SNPs are closely entangled with the MCC rate estimate, and both cannot be estimated at the same time, an iterative estimation method is adopted, whereby the estimate of each one is updated with the present estimate of the other,and this process is repeated until convergence. At each stage, the MCC rate is first estimated using all SNPs labeled as PM, and then the haplotype status for each SNP is estimated using the new MCC rate estimate. This process is repeated until the difference between two adjacent estimates of MCC rate does not exceed a threshold (set to be 0.1 or 0.05). The estimate in the final iteration is used as the final estimate of MCC rate and the corresponding haplotype states are adopted as the final labels of haplotype status. If the MCC rate estimates do not converge after 10 rounds of iteration, the average of the 10 MCC rate estimates is then taken as the final estimate, and the haplotype status is updated using this estimate.

The sample of culture medium with an abnormal initial or final estimated MCC rate (>0.75 or <-0.5) is considered to be of extremely low quality. Thus, the sample is excluded and classified as “Rule out”.

Step 5: Calculating the likelihood of observing the SNP data within regions adjacent to the disease-causing mutation site, and combining the likelihood at each single SNP and the recombination probabilities.

First, if the disease-causing mutation site is on the paternal chromosome, all SNPs whose final haplotype status is labeled as P or PM and where the paternal genotype is heterozygous are selected for the calculation of likelihood. If the disease-causing mutation site is on the maternal chromosome, all SNPs whose final haplotype status is labeled M or PM and where the maternal genotype is heterozygous are selected for the calculation of likelihood. The disease-causing mutation site is still denoted as the 0-th SNP. All retained SNPs downstream of the 0-th SNP have ordinal numbers 1, 2, ..., $N_2$, while the upstream SNPs have ordinal numbers −1, −2,...,$-N_1$. For the i-th SNP, it is denoted

$$P_i = \begin{cases} 1 & \text{If the embryonic chromosome inherited from the father was actually from the grandfather} \\ 2 & \text{If the embryonic chromosome inherited from the father was actually from the grandmother} \end{cases}$$

$$M_i = \begin{cases} 1 & \text{If the embryonic chromosome inherited from the mother was actually from the grandfather} \\ 2 & \text{If the embryonic chromosome inherited from the mother was actually from the grandmother} \end{cases}$$

$AD_i$ is denoted as the Allele Depth (AD) at the i-th SNP and $AD_{i:j} = \{AD_i, AD_{i+1}, ..., AD_{i+j}\}$ for i < j. Given the genotype and haplotype labels at the disease-causing mutation site, the recombination and sequencing data in the upstream and downstream are conditionally independent, and the upstream and downstream analysis are dealt with separately. Here, formulating the recursion for the downstream region is further described. The probability that the parental embryonic allele is inherited from the grandfather or grandmother and the maternal embryonic allele is inherited from grandfather or grandmother is calculated, conditioned on the observation of all detected AD values and the inferred haplotype status for SNPs within the adjacent region. Specifically, the goal is to calculate

$$\mathrm{P}\left(P_0 = i, M_0 = j | AD_{0:N_2}, H_{0:N_2}\right), \text{ for i, j=1, 2.}$$

The following Bayesian formula is utilized

$$\mathrm{P}\left(P_0 = i, M_0 = j | AD_{0:N_2}, H_{0:N_2}\right) \propto \mathrm{P}(P_0 = i, M_0 = j)\mathrm{P}\left(AD_{0:N_2} | P_0 = i, M_0 = j, H_{0:N_2}\right)$$

with non-informative prior:

$$\mathrm{P}(P_0 = i, M_0 = j) = \frac{1}{4}, \text{ for i, j=1,2.}$$

The next step is to calculate $\mathrm{P}\left(AD_{0:N_2} | P_0 = i, M_0 = j, H_{0:N_2}\right)$.

$$\mathrm{P}\left(AD_{n:N_2} | P_n = i, M_n = j, H_{n:N_2}\right)$$
$$= L_{ij}^{(n)} \sum_{k,l=1}^{2} \mathrm{P}\left(AD_{n+1:N_2} | P_{n+1} = k, M_{n+1} = l, H_{n+1:N_2}\right) \cdot P(P_{n+1} = k, M_{n+1} = l | P_n = i, M_n = j)$$

The single-SNP likelihood is $L_{ij}^{(n)} = \mathrm{P}(AD_n | P_n = i, M_n = j, H_n)$ and the initial value is $L_{ij}^{(N_2)}$. $P(F_{n+1} = k, M_{n+1} = l | P_n = i, M_n = j)$ is the recombination probability.

The procedure described above reduces the calculation of the likelihood function to the calculation of the recombination probability and the single-SNP likelihood. The estimated recombination probability between any two positions on the human genome can be obtained from a high-precision database of recombination rates, such as [11,12].

Step 6): Determining whether the embryo carries the disease-causing haplotype, and providing a level of confidence.

After calculating the likelihood, it is determined which of the father's two haplotypes and which of the mother's two haplotypes is inherited by the embryo by calculating Log Likelihood Ratio (LLR). If the disease-causing mutation site is on the paternal chromosome, the following is defined

$$LLR = \log\left(\frac{\sum_{j=1}^{2} \mathbb{P}(AD_{0:N} | P_0 = 1, M_0 = j)}{\sum_{j=1}^{2} \mathbb{P}(AD_{0:N} | P_0 = 2, M_0 = j)}\right),$$

otherwise we define

$$LLR = \log\left(\frac{\sum_{j=1}^{2} \mathbb{P}(AD_{0:N} | P_0 = j, M_0 = 1)}{\sum_{j=1}^{2} \mathbb{P}(AD_{0:N} | P_0 = j, M_0 = 2)}\right).$$

A positive LLR indicates that the data considered favors that the allele is inherited from the father of the disease-carrying parent. Otherwise, the data supports that it is inherited from the mother of the disease-carrying parent.

We recognize that the ideal number of SNPs to use for the calculation of likelihood cannot be determined in advance. If a fixed number of SNPs is used for the calculation, or if all SNPs within a fixed physical distance to the disease-causing mutation site is used for the calculation, it cannot be determined with certainty ahead of time whether the information implied by these SNPs will be sufficient to support conclusions, and to what extent the likelihood varies with the number of SNPs used.

Therefore, SNPs are successively added into the calculation of likelihood and for each $N_2 = 1, 2, ...$, the likelihood is calculated based on which parent's chromosome the disease-causing mutation site is on. The LLR curve is then plotted against the physical distance of the SNP to the disease-causing mutation site. Typically, when the number of used SNPs is small, the data itself contains insufficient information about whether the embryo carries the disease-carrying allele, so the absolute value of the log-likelihood ratio is expected to be small and the curve will be close to the horizontal axis. As the number of used SNPs increases, more and more information is taken into consideration, and if the internal consistency of the data is good, the curve will plateau with a sufficiently large absolute value. To make the log-likelihood curve more stable, the log-likelihood change for each pair of adjacent SNPs is limited to no more than 5.

Finally, when the number of used SNPs is large enough to determine whether the embryo carries the disease-carrying allele, adding new SNPs causes only very small changes in the log-likelihood ratio. Therefore, when the vertical coordinates of the curve become stable, this can be used as a criterion to determine whether the embryo carries the disease-carrying allele, and its absolute value serves as a measure for the confidence level. If either upstream or downstream region has fewer than 10 SNPs, this region is ignored and the confidence level according to the curve on the other side is determined. This can happen when the disease-causing mutation site is at either end of the chromosome, or near the centromere.

The confidence level is classified into four categories: High Confidence, Moderate Confidence, Low Confidence, and Undetermined. The confidence levels are categorized based on the following standards:

High Confidence

+ The average steady value, which is the average of upstream and downstream steady values when sufficient SNP data is provided, is ≥ 5, and there are no differing signs.

+ At least one steady value ≥ 1.

Moderate Confidence

+ The maximum absolute value of the likelihood on the curve with differing signs from the steady value is < 5, while the average steady value is ≥ 3.

+ At least one steady value ≥ 1.

Low Confidence

+ All other samples that are not categorized as Highly Confident, Moderately Confident, or Undetermined

Undetermined

1. Steady values have differing signs.

2. Either side has a steady value < 0.5, indicating low confidence evidence.

3. - The maximum absolute value of the likelihood on the curve with differing signs from the steady value is > 20.

**Rule Out** conditions:

- Error rate ≥ 0.2.
- Low GQ (GQ<=3) rate ≥ 0.4.
- Original MCC rate or final MCC rate:
    * ≥ 0.75 or ≤ -0.5 for cultural media.
    * ≥ 0.85 or ≤ -0.7 for cultural media of discarded embryos.
- SNP density ≤ 0.001.

Following the above protocol, the genotype and which allele is inherited from parents at any single SNP can be determined. This information can then be used to execute the disease-carrying analysis for monogenic diseases.

For X-linked recessive disease, we first use CNV to determine the gender of the embryo, and then if the disease-carrying mutation site is on the X chromosome, we set the haplotype state of all the SNP as “M” if the embryo is male.

### 3. Whole Genome Construction

In DNA samples prepared from spent embryo culture medium (SCM), many genomic regions contained information from only one parent. This allelic dropout can occur from single-cell whole genome amplification bias or large fragment loss of haplotype (Extended Data Fig. 5). The conventional diploid assumption of the genome therefore cannot be applied to haplotyping and genotype imputation of SCM samples. Therefore, we developed a Pedigree-Population-based Imputation with Haploid Assumption (PPIHA) to reconstruct whole genomes from SCM. Specifically, we combined pre-phased parental haplotype information and selected informative SNPs where at least one parental origin haplotype could be confidently phased to create the haplotype scaffold of the embryo from SCM data (Extended Data Fig. 9). Initially, the SCM genotype determination is made independent of information about the parents. For example, at a given locus the SCM genotype may initially be called "BB". However, based on the pre-phased parental haplotypes, the paternal genotype was AB (allele A comes from haplotype P1 and allele B from P2), and the maternal genotype was AA. With this information and based on the rules of Mendelian inheritance it can be inferred that the B allele in SCM is derived from haplotype P2, and that the Maternal haplotype cannot be determined at this site due to allele dropout or large fragment loss in the SCM sample (Extended Data Fig. 5). And likewise, for a locus with maternal, paternal and SCM genotypes being AB, AA and BB, respectively, the maternal haplotype of SCM could be inferred while paternally genetic information was unavailable. After examining all such kinds of informative variants from the sequenced SCM data, we constructed a sparse haplotype scaffold with a single parent of the embryo (Extended Data Fig. 8).

Paternal or maternal haplotypes in the scaffold were first corrected if continuous variants of the opposite haplotype were observed on both nearby sides. This correction was applied when the probability of a double crossover between the nearest variants with the opposite haplotype, calculated as the square of their genetic distances multiplied by 0.0001, was less than 0.005 for the maternal haplotype or 0.001 for the paternal haplotype, according to the chromosome interference theory [14]. Then, with parental haplotype support and the Hidden-Markov-Model strategy, paternal and maternal haplotypes of other variants were independently phased based on a pre-determined haplotype scaffold of SCM (Extended Data Fig. 8). We assumed that the state (P1 or P2 for paternal haplotypes and M1 or M2 for maternal haplotypes) at variant t could be inferred by SNPs in the scaffold from t-5 to t+5. The transition probability between two SNPs was considered equivalent to their genetic distance multiplied by 0.01. Joint probabilities of the two states of paternal or maternal haplotypes at variant t were calculated using a forward-backward algorithm based on the adjoining ten variants (from t-5 to t+5). The haplotype of variant t was then assigned to the state with the higher probability. If the joint probabilities of the two states (P1/P2 or M1/M2) for a variant were equal, phasing of that variant failed. By organizing the haplotype states of all variants according to their chromosomal positions, the pedigree-based haplotypes of SCMs were reconstructed. Subsequently, the genotypes of SCM were imputed with the genome information of the parents (Extended Data Fig. 8).

Following pedigree-based genome reconstruction, population-based genotype imputation was conducted using the 1000 Genomes haplotype reference panel with Minimac4 [15], resulting in the reconstruction of the whole genome information of the embryo from the SCM samples (Extended Data Fig. 8).

We employed a PRS model of T2D including 128 SNPs from a previous study [16]. To validate the model and determine the PRS percentile (indicating relative disease risk) in the UK Biobank (UKB) cohort, SNPs whose genotype was not available or failed to be imputed in UKB cohort were excluded, leaving a total of 114 SNPs. We then directly used the effect sizes from the original study to calculate the PRS, employing the following formula:

$$\text{PRS} = \sum_{i=1}^{114} \beta_i \times G_i$$

where $G_i$ is the allelic dosage and $\beta_i$ is the effect size of SNP $i$.

Using the T2D model and control information in the UKB cohort, the PRS percentile for T2D was obtained to evaluate the polygenic risk of T2D for each individual.

References：

#### Table S1 Amplification Bias according to DNA fragment length.

| Method | Reads detected in regions 1/Reads detected in regions 2 | Normalized |
|---|---|---|
| Bulk (no amplification) | 4.4 | 1 |
| MDA | 2,193.9 | 504.3 |
| MALBAC | 25,900.7 | 5,954.2 |
| Improved LIANTI | 22.2 | 5.1 |
| Region 1 length=1304bp; Region 2 length=186bp | | |

MDA, MALBAC, and Improved LIANTI were used to amplify two different DNA regions of different length with the equal molarity. After amplification, the number of sequencing reads mapped to each of the regions was normalized with the "bulk" sample (no amplification). The MALBAC amplification method strongly favored amplification of longer fragments, while the improved LIANTI exhibited the least template length bias.

#### Table S2. Analysis of BJ15 and BJ17 culture medium DNA amplification with three methods.

| Sample | Reads | Map% | Coverage % | Multi_ map% | Dup% | SNP_unflt | SNP_flt | Flt% | Common_pos | Overlap % | Sensitivity % | Accuracy % | Dp_ median | ts/tv | rs | rs% |
|---|---|---|---|---|---|---|---|---|---|---|---|---|---|---|---|---|
| MALBAC_BJ15 | 190,951,730 | 94 | 35 | 2 | 18 | 2,719,783 | 1,739,603 | 64 | 472,155 | 27 | 14 | 99 | 15 | 1 | 626,392 | 36 |
| MALBAC_BJ17 | 149,260,612 | 99 | 40 | 2 | 12 | 2,896,927 | 1,810,515 | 62 | 501,425 | 28 | 15 | 100 | 13 | 1 | 660,780 | 36 |
| MDA_BJ15 | 241,991,652 | 100 | 33 | 12 | 12 | 1,057,965 | 581,289 | 55 | 528,512 | 91 | 16 | 100 | 20 | 2 | 553,195 | 95 |
| MDA_BJ17 | 236,484,214 | 99 | 56 | 12 | 10 | 1,741,749 | 1,015,977 | 58 | 932,702 | 92 | 28 | 100 | 15 | 2 | 972,572 | 96 |
| LIANTI_BJ15_Improved | 177,107,562 | 99 | 52 | 4 | 24 | 1,629,861 | 779,484 | 48 | 691,622 | 89 | 21 | 100 | 12 | 2 | 729,134 | 94 |
| LIANTI_BJ17_Improved | 167,665,852 | 99 | 61 | 3 | 17 | 1,858,050 | 979,819 | 53 | 883,957 | 90 | 26 | 100 | 11 | 2 | 927,208 | 95 |
| Bulk_BJ_Cell | 907,593,012 | 97 | 92 | 2 | 6 | 4,087,217 | 3,340,555 | 82 | 3,340,555 | 100 | 100 | 100 | 36 | 2 | 3,334,025 | 100 |

In this study, Genome coverage (Column 4, yellow background) and the number of reference SNPs detected (Column 16, red background) were chosen as key metrics. For both BJ15 (Shorter Overall DNA fragments, Fig S6) and BJ17 (Longer Overall DNA fragments DNA, Fig S6), improved LIANTI was superior to the other two amplification methods in terms of genome coverage and number of SNPS, even with fewer reads. Therefore, we chose the improved LIANTI method to amplify the cell-free DNA from spent culture medium.

**Table S3. Maternal cell contamination rate of SCM samples in autosomal dominant and autosomal recessive familly**

| Sample | orimcc | finalmcc |
|---|---:|---:|
| 1T-SCM1 | -0.075 | 0.01 |
| 1T-SCM2 | 0.041 | 0.01 |
| 1T-SCM3 | 0.878 | 0.878 |
| 1T-SCM4 | 0.401 | 0.781 |
| 1T-SCM5 | 0.425 | 0.425 |
| 1T-SCM6 | 0.317 | 0.317 |
| 2T-SCM1 | 0.471 | 0.188 |
| 2T-SCM2 | 0.274 | 0.086 |
| 2T-SCM3 | 0.491 | 0.491 |
| 2T-SCM4 | 0.46 | 0.01 |
| 2T-SCM5 | 0.397 | 0.0625 |
| 2T-SCM6 | 0.027 | 0.036 |
| 2T-SCM7 | -0.309 | 0.098 |
| 2T-SCM8 | -0.031 | 0.034 |
| 3D-SCM1 | 0.131 | 0.01 |
| 3D-SCM2 | 0.906 | 0.761 |
| 3D-SCM3 | -0.156 | 0.01 |
| 3T-SCM1 | 0.186 | 0.013 |
| 3T-SCM2 | 0.554 | 0.141 |
| 3T-SCM3 | 0.051 | 0.01 |
| 3T-SCM4 | -0.148 | 0.01 |
| 3T-SCM5 | 0.063 | 0.03 |
| 4D-SCM1 | 0.116 | 0.208 |
| 4D-SCM2 | 0.757 | 0.01 |
| 4D-SCM3 | -0.133 | 0.01 |
| 4D-SCM4 | -0.027 | 0.053 |
| 4T-SCM1 | 0.449 | 0.01 |
| 4T-SCM2 | 0.092 | 0.365 |
| 4T-SCM3 | 0.004 | 0.209 |
| 4T-SCM4 | 0.792 | 0.091 |
| 4T-SCM5 | -0.088 | 0.158 |
| 4T-SCM6 | 0.14 | 0.194 |
| 4T-SCM7 | 0.207 | 0.128 |
| 5D-SCM1 | 0.284 | 0.508 |
| 5D-SCM2 | 0.036 | 0.01 |
| 5T-SCM1 | 0.04 | 0.338 |
| 5T-SCM2 | 0.965 | 0.943 |
| 6D-SCM1 | 0.027 | 0.01 |
| 6D-SCM2 | -0.086 | 0.195 |
| 6T-SCM1 | -0.217 | 0.01 |
| 6T-SCM2 | -0.054 | 0.01 |
| 7D-SCM1 | -0.233 | 0.125 |
| 7T-SCM1 | 0.375 | 0.01 |
| 7T-SCM2 | 0.006 | 0.14 |
| 8T-SCM1 | 0.15 | 0.161 |
| 8T-SCM2 | -0.28 | 0.01 |
| 8T-SCM3 | -0.1149 | 0.01 |
| 8T-SCM4 | -0.551 | 0.01 |
| 8T-SCM5 | 0.934 | 0.877 |

| | | |
|---|---|---|
| 9D-SCM1 | 0.027 | 0.01 |
| 9D-SCM2 | 0.028 | 0.135 |
| 9D-SCM3 | 0.035 | 0.01 |
| 9D-SCM4 | 0.335 | 0.078 |
| 9T-SCM1 | 0.91 | 0.01 |
| 9T-SCM2 | 0.146 | 0.2 |
| 9T-SCM3 | -0.191 | 0.01 |
| 9T-SCM4 | 0.655 | 0.01 |
| 9T-SCM5 | 0.063 | 0.037 |
| 9T-SCM6 | -0.094 | 0.01 |
| 9T-SCM7 | 0.116 | 0.056 |
| 9T-SCM8 | 0.349 | 0.207 |
| 9T-SCM9 | 0.053 | 0.01 |
| 10D-SCM1 | 0.078 | 0.01 |
| 10D-SCM2 | -0.444 | 0.01 |
| 10T-SCM1 | 0.0002 | 0.153 |
| 10T-SCM2 | 0.767 | 0.01 |
| 10T-SCM3 | 0.6566 | 0.2075 |
| 11D-SCM1 | -0.051 | 0.01 |
| 11D-SCM2 | 0.425 | 0.425 |
| 11D-SCM3 | 0.187 | 0.01 |
| 11T-SCM1 | -0.032 | 0.01 |
| 11T-SCM2 | -0.005 | 0.01 |
| 11T-SCM3 | -0.041 | 0.015 |
| 11T-SCM4 | -0.018 | 0.01 |
| 11T-SCM5 | 0.065 | 0.052 |
| 12D-SCM1 | -0.086 | 0.01 |
| 12D-SCM2 | -0.032 | 0.01 |
| 12D-SCM3 | 0.416 | 0.089 |
| 12D-SCM4 | 0.997 | 0.997 |
| 12T-SCM1 | 0.056 | 0.044 |
| 15D-SCM1 | 0.0199 | 0.0343 |
| 15T-SCM1 | 0.9988 | 0.9614 |
| 15T-SCM2 | 0.0312 | 0.0100 |
| 15T-SCM3 | 0.0274 | 0.0163 |
| 15T-SCM4 | 0.1205 | 0.0356 |
| 16D-SCM1 | 0.0827 | 0.0548 |
| 16D-SCM2 | 0.0511 | 0.0100 |
| 16D-SCM3 | 0.0809 | 0.0485 |
| 16T-SCM1 | 0.2961 | 0.0828 |
| 16T-SCM10 | 0.3306 | 0.0100 |
| 16T-SCM11 | 0.2780 | 0.3161 |
| 16T-SCM12 | -0.0763 | 0.0028 |
| 16T-SCM2 | 0.1259 | 0.1590 |
| 16T-SCM3 | -0.5350 | 0.0100 |
| 16T-SCM4 | 0.0250 | 0.0100 |
| 16T-SCM5 | 0.6627 | 0.4000 |
| 16T-SCM6 | -0.0568 | 0.0588 |
| 16T-SCM7 | 0.1313 | 0.0100 |
| 16T-SCM8 | 0.0315 | 0.0258 |
| 16T-SCM9 | 0.1273 | 0.0288 |
| 17D-SCM1 | 0.0885 | 0.1206 |
| 17D-SCM2 | -0.0440 | 0.0100 |
| 17D-SCM3 | -0.1664 | 0.0100 |

| | | |
|---|---|---|
| 17T-SCM1 | 0.9143 | 0.9143 |
| 17T-SCM2 | 0.0798 | 0.2768 |
| 17T-SCM3 | -0.1202 | 0.0100 |
| 17T-SCM4 | 0.0591 | 0.1462 |
| 17T-SCM5 | 0.1110 | 0.0100 |
| 17T-SCM6 | -0.3443 | 0.0100 |
| 17T-SCM7 | 0.0474 | 0.0100 |
| 17T-SCM8 | 0.1554 | 0.0100 |
| 18D-SCM1 | 0.0011 | 0.0434 |
| 18D-SCM2 | -0.0704 | 0.1015 |
| 18D-SCM3 | -0.0846 | 0.0415 |
| 18D-SCM4 | 0.2052 | 0.0753 |
| 18D-SCM5 | 0.0428 | 0.0308 |
| 18D-SCM6 | 0.1003 | 0.0601 |
| 18D-SCM7 | 0.0384 | 0.0100 |
| 18D-SCM8 | 0.1524 | 0.0616 |
| 18T-SCM1 | 0.9965 | 0.9153 |
| 18T-SCM2 | -0.0294 | 0.0245 |
| 18T-SCM3 | -0.0071 | 0.0005 |
| 18T-SCM4 | 0.0923 | 0.0430 |
| 18T-SCM5 | 0.2519 | 0.3374 |
| 18T-SCM6 | 0.0569 | 0.0262 |
| 18T-SCM7 | 0.9888 | 0.9924 |
| 18T-SCM8 | -0.0124 | 0.0074 |
| 19D-SCM1 | 0.4677 | 0.0100 |
| 19D-SCM2 | -0.0589 | 0.0198 |
| 19T-SCM1 | 0.9983 | 0.9983 |
| 19T-SCM2 | 0.1609 | 0.0100 |
| 19T-SCM3 | 0.5560 | 0.1217 |
| 20T-SCM1 | 0.3003 | 0.2368 |
| 20T-SCM2 | 0.0000 | 0.0100 |
| 21T-SCM1 | 0.1802 | 0.0100 |
| 21T-SCM2 | 0.1858 | 0.0100 |
| 21T-SCM3 | -0.0339 | 0.0100 |
| 21T-SCM4 | 0.0293 | 0.1710 |
| 22T-SCM1 | 0.1944 | 0.0100 |
| 22T-SCM2 | 0.0188 | 0.0100 |
| 22T-SCM3 | 0.2617 | 0.0100 |
| 22T-SCM4 | 0.1750 | 0.0365 |
| 23D-SCM1 | 0.3022 | 0.1615 |
| 23D-SCM2 | -0.1408 | 0.0052 |
| 23D-SCM3 | 0.5505 | 0.2706 |
| 23D-SCM4 | 0.2716 | 0.0994 |
| 23D-SCM5 | -0.0172 | 0.0100 |
| 23T-SCM1 | 0.1857 | 0.0100 |
| 23T-SCM2 | 0.3495 | 0.3495 |
| 24D-SCM1 | -0.0022 | 0.0100 |
| 24D-SCM2 | 0.1946 | 0.0100 |
| 24D-SCM3 | -0.4091 | 0.0100 |
| 24D-SCM4 | 0.3134 | 0.3134 |
| 24D-SCM5 | 0.0849 | 0.0100 |
| 24T-SCM1 | 0.2076 | 0.0100 |
| 24T-SCM2 | 0.2555 | 0.6059 |
| 24T-SCM3 | 0.0033 | 0.0100 |

| | | |
|---|---|---|
| 25T-SCM1 | 0.0796 | 0.0028 |
| 25T-SCM10 | 0.1453 | 0.0100 |
| 25T-SCM2 | 0.0928 | 0.0100 |
| 25T-SCM3 | 0.2494 | 0.0100 |
| 25T-SCM4 | -0.0546 | 0.0091 |
| 25T-SCM5 | 0.2339 | 0.0099 |
| 25T-SCM6 | 0.4981 | 0.5161 |
| 25T-SCM7 | 0.9845 | 0.5402 |
| 25T-SCM8 | 0.5421 | 0.1765 |
| 25T-SCM9 | 0.9818 | 0.9818 |
| 26T-SCM1 | -0.2742 | 0.0100 |
| 26T-SCM2 | -0.0334 | 0.0100 |
| 26T-SCM3 | 0.0600 | 0.0928 |
| 26T-SCM4 | 0.4401 | 0.0797 |
| 27T-SCM1 | 0.2637 | 0.0385 |
| 27T-SCM2 | 0.6847 | 0.0100 |
| 27T-SCM3 | 0.1898 | 0.0100 |
| 27T-SCM4 | -0.0577 | 0.3235 |
| 27T-SCM5 | -0.0718 | 0.0100 |
| 28T-SCM1 | -0.1649 | 0.0005 |
| 28T-SCM2 | 0.2053 | 0.0100 |
| 29T-SCM1 | 0.0870 | 0.0288 |
| 29T-SCM2 | 0.5420 | 0.0812 |
| 29T-SCM3 | 0.9027 | 0.0100 |
| 29T-SCM4 | -0.1083 | 0.1036 |

**Table S4. Number of paired samples used for noninvasive preimplantation genetic testing for polygenic disorders and polygenic risk score calculation.**

| Family | TE biopsy samples | Discarded embryos | SCM samples for whole genome reconstruction | SCM samples for niPGT-P | Amniotic fluid | Amniotic fluid with QC-passed SCM samples |
|---|---|---|---|---|---|---|
| 1 | 6 | 0 | 5 | 1 | 1 | 1 |
| 2 | 8 | 0 | 7 | 5 | 0 | 0 |
| 3 | 5 | 2 | 5 | 4 | 1 | 1 |
| 4 | 7 | 4 | 1 | 0 | 1 | 0 |
| 5 | 2 | 1 | 0 | 0 | 0 | 0 |
| 6 | 2 | 1 | 0 | 0 | 0 | 0 |
| 7 | 2 | 2 | 2 | 2 | 0 | 0 |
| 8 | 5 | 0 | 2 | 1 | 0 | 0 |
| 9 | 9 | 4 | 4 | 4 | 0 | 0 |
| 10 | 3 | 2 | 4 | 2 | 1 | 0 |
| 11 | 5 | 3 | 4 | 2 | 0 | 0 |
| 12 | 1 | 4 | 2 | 0 | 0 | 0 |
| 13 | 6 | 0 | 5 | 3 | 0 | 0 |
| 14 | 3 | 0 | 2 | 1 | 0 | 0 |
| Sum | 64 | 23 | 43 | 25 | 4 | 2 |

**Table S5. Summary Statistics and niPGT-M results for all samples.**

| Family | Chrom | Pos | Disease name | Parent | Sample | Coverage | Rate of GQ <= 3 | err_rate (20M) | niPGT-M | PGT-M | Classification |
|---|---|---|---|---|---|---|---|---|---|---|---|
| 1 | 14 | 92524896 | SCA3, expansion of CAG repeats | M | 1T-SCM1 | 39.706 | 0.143 | 0 | - | - | High |
| | | | | | 1T-SCM2 | 66.645 | 0.185 | 0 | + | + | High |
| | | | | | 1T-SCM3 | 23.585 | 0.113 | 0 | / | - | Rule Out |
| | | | | | 1T-SCM4 | 32.024 | 0.128 | 0 | / | - | Rule Out |
| | | | | | 1T-SCM5 | 21.097 | 0.15 | 0.005 | / | + | Rule Out |
| | | | | | 1T-SCM6 | 5.626 | 0.144 | 0 | / | + | Rule Out |
| 2 | 15 | 48829901 | FBN1, C.643C>T | P | 2T-SCM1 | 16.337 | 0.019 | 0.081 | / | - | Undetermined |
| | | | | | 2T-SCM2 | 13.524 | 0.021 | 0.041 | / | - | Undetermined |
| | | | | | 2T-SCM3 | 28.995 | 0.033 | 0.084 | + | + | High |
| | | | | | 2T-SCM4 | 29.644 | 0.04 | 0.067 | + | + | High |
| | | | | | 2T-SCM5 | 42.519 | 0.058 | 0.043 | + | + | High |
| | | | | | 2T-SCM6 | 24.884 | 0.031 | 0.073 | + | + | High |
| | | | | | 2T-SCM7 | 55.826 | 0.077 | 0.047 | - | - | Moderate |
| | | | | | 2T-SCM8 | 51.607 | 0.083 | 0.023 | - | - | High |
| 3 | 16 | 2155477 | PKD1, c.7864_2A>G | P | 3T-SCM1 | 44.033 | 0.144 | 0.006 | - | - | High |
| | | | | | 3T-SCM2 | 37.65 | 0.167 | 0.007 | - | - | Low |
| | | | | | 3T-SCM3 | 59.288 | 0.154 | 0.008 | + | + | Low |
| | | | | | 3T-SCM4 | 42.302 | 0.136 | 0.008 | + | + | High |
| | | | | | 3T-SCM5 | 80.926 | 0.129 | 0.008 | - | - | High |
| | | | | | 3D-SCM1 | 56.692 | 0.129 | 0.007 | - | - | High |
| | | | | | 3D-SCM2 | 35.703 | 0.157 | 0.005 | / | - | Rule Out |
| | | | | | 3D-SCM3 | 33.539 | 0.149 | 0.007 | + | + | High |
| 4 | 7 | 42002168 | GLI3, C.1880-1881delAT | M | 4T-SCM1 | 8.439 | 0.462 | 0.149 | / | - | Rule Out |
| | | | | | 4T-SCM2 | 30.293 | 0.249 | 0.057 | - | - | Moderate |
| | | | | | 4T-SCM3 | 29.969 | 0.325 | 0.1 | + | + | High |
| | | | | | 4T-SCM4 | 2.813 | 0.487 | 0.125 | / | - | Rule Out |
| | | | | | 4T-SCM5 | 25.208 | 0.341 | 0.086 | + | + | Moderate |
| | | | | | 4T-SCM6 | 27.048 | 0.387 | 0.056 | + | + | High |
| | | | | | 4T-SCM7 | 26.615 | 0.343 | 0.104 | - | - | Moderate |
| | | | | | 4D-SCM1 | 40.571 | 0.318 | 0.085 | + | + | Moderate |
| | | | | | 4D-SCM2 | 11.035 | 0.344 | 0.176 | + | + | Moderate |
| | | | | | 4D-SCM3 | 15.038 | 0.282 | 0.055 | - | - | High |
| | | | | | 4D-SCM4 | 36.136 | 0.324 | 0.048 | - | - | Moderate |

| | | | | | | | | | | | |
|---|---|---|---|---|---|---|---|---|---|---|---|
| 5 | 16 | 2160372 | PKD1, c.4795dupT | P | 5T-SCM1 | 19.907 | 0.153 | 0.006 | + | + | High |
| | | | | | 5T-SCM2 | 28.779 | 0.167 | 0.005 | / | + | Rule Out |
| | | | | | 5D-SCM1 | 19.15 | 0.186 | 0.006 | + | + | Moderate |
| | | | | | 5D-SCM2 | 24.343 | 0.168 | 0.004 | - | - | Moderate |
| 6 | 14 | 92524896 | SCA3, expansion of CAG repeats | P | 6T-SCM1 | 25.966 | 0.157 | 0.017 | - | - | High |
| | | | | | 6T-SCM2 | 38.624 | 0.114 | 0.003 | + | + | Low |
| | | | | | 6D-SCM1 | 31.267 | 0.117 | 0.006 | - | - | High |
| | | | | | 6D-SCM2 | 49.227 | 0.163 | 0.008 | - | - | Low |
| 7 | 16 | 2153695 | PKD1, C.8363C>A | P | 7T-SCM1 | 28.887 | 0.14 | 0.008 | + | + | Low |
| | | | | | 7T-SCM2 | 37.975 | 0.125 | 0.008 | + | + | Low |
| | | | | | 7D-SCM1 | 44.791 | 0.14 | 0.006 | - | - | High |
| 8 | 1 | 45974647 | MMACHC, C.609G>A | P | 8T-SCM1 | 57.341 | 0.126 | 0.022 | + | + | High |
| | | | | | 8T-SCM2 | 57.341 | 0.232 | 0.035 | + | + | High |
| | | | | | 8T-SCM3 | 29.103 | 0.2496 | 0.0424 | - | - | Moderate |
| | | | | | 8T-SCM4 | 26.074 | 0.228 | 0.083 | / | + | Rule out |
| | | | | | 8T-SCM5 | 43.709 | 0.109 | 0.009 | / | + | Rule Out |
| | 1 | 45974647 | MMACHC, C.609G>A | M | 8T-SCM1 | 57.341 | 0.126 | 0.022 | + | + | High |
| | | | | | 8T-SCM2 | 57.341 | 0.232 | 0.035 | + | + | Moderate |
| | | | | | 8T-SCM3 | 29.103 | 0.2496 | 0.0424 | / | + | Undetermined |
| | | | | | 8T-SCM4 | 26.074 | 0.228 | 0.083 | / | - | Rule Out |
| | | | | | 8T-SCM5 | 43.709 | 0.109 | 0.009 | / | - | Rule Out |

| | | | | | | | | | | | |
|---|---|---|---|---|---|---|---|---|---|---|---|
| 9 | 10 | 14987109 | DCLRE1C, C.241C>T | P | 9T-SCM1 | 1.406 | 0.233 | 0.263 | / | - | Rule Out |
| | | | | | 9T-SCM2 | 8.547 | 0.274 | 0.071 | + | + | Moderate |
| | | | | | 9T-SCM3 | 11.793 | 0.168 | 0.0175 | + | + | High |
| | | | | | 9T-SCM4 | 6.275 | 0.453 | 0.113 | / | + | Rule Out |
| | | | | | 9T-SCM5 | 82.874 | 0.103 | 0.012 | - | - | High |
| | | | | | 9T-SCM6 | 56.151 | 0.161 | 0.017 | + | + | High |
| | | | | | 9T-SCM7 | 39.381 | 0.168 | 0.047 | - | - | High |
| | | | | | 9T-SCM8 | 40.247 | 0.214 | 0.024 | + | + | High |
| | | | | | 9T-SCM9 | 42.411 | 0.15 | 0.041 | - | - | Moderate |
| | | | | | 9D-SCM1 | 46.738 | 0.117 | 0.035 | - | - | Low |
| | | | | | 9D-SCM2 | 47.387 | 0.318 | 0.037 | / | - | Moderate |
| | | | | | 9D-SCM3 | 45.981 | 0.113 | 0.013 | - | - | Low |
| | | | | | 9D-SCM4 | 62.101 | 0.139 | 0.017 | / | - | Undetermined |
| | 10 | 14950816 | DCLRE1C, C.1372G>T | M | 9T-SCM1 | 1.406 | 0.233 | 0.263 | / | + | Rule Out |
| | | | | | 9T-SCM2 | 8.547 | 0.274 | 0.072 | + | + | Low |
| | | | | | 9T-SCM3 | 11.793 | 0.168 | 0.0175 | / | - | Undetermined |
| | | | | | 9T-SCM4 | 6.275 | 0.453 | 0.113 | / | + | Rule Out |
| | | | | | 9T-SCM5 | 82.874 | 0.103 | 0.012 | + | + | High |
| | | | | | 9T-SCM6 | 56.151 | 0.161 | 0.017 | - | - | Moderate |
| | | | | | 9T-SCM7 | 39.381 | 0.168 | 0.047 | - | - | Moderate |
| | | | | | 9T-SCM8 | 40.247 | 0.214 | 0.024 | + | + | High |
| | | | | | 9T-SCM9 | 42.411 | 0.15 | 0.041 | - | - | High |
| | | | | | 9D-SCM1 | 46.738 | 0.117 | 0.035 | + | + | High |
| | | | | | 9D-SCM2 | 47.387 | 0.318 | 0.036 | + | + | High |
| | | | | | 9D-SCM3 | 45.981 | 0.113 | 0.013 | - | - | High |
| | | | | | 9D-SCM4 | 62.101 | 0.139 | 0.017 | - | - | Low |
| 10 | 1 | 45974001 | MMACHC, c.394C>T | P | 10T-SCM1 | 41.761 | 0.118 | 0.014 | + | + | High |
| | | | | | 10T-SCM2 | 26.074 | 0.113 | 0.017 | / | - | Rule Out |
| | | | | | 10T-SCM3 | 53.446 | 0.1226 | 0.0238 | / | - | Undetermined |
| | | | | | 10D-SCM1 | 63.616 | 0.135 | 0.025 | - | - | Moderate |
| | | | | | 10D-SCM2 | 64.049 | 0.114 | 0.027 | - | - | High |
| | 1 | 45974647 | MMACHC, c.609G>A | M | 10T-SCM1 | 41.761 | 0.118 | 0.014 | - | - | High |
| | | | | | 10T-SCM2 | 26.074 | 0.113 | 0.017 | / | - | Rule Out |
| | | | | | 10T-SCM3 | 53.446 | 0.1226 | 0.0238 | + | + | Low |
| | | | | | 10D-SCM1 | 63.616 | 0.135 | 0.025 | - | - | Moderate |
| | | | | | 10D-SCM2 | 64.049 | 0.114 | 0.027 | + | + | High |

| | | | | | | | | | | | |
|---|---|---|---|---|---|---|---|---|---|---|---|
| 11 | 1 | 45974647 | MMACHC, c.80A>G | P | 11T-SCM1 | 14.497 | 0.137 | 0.019 | + | + | Moderate |
| | | | | | 11T-SCM2 | 50.849 | 0.14 | 0.014 | / | - | Undetermined |
| | | | | | 11T-SCM3 | 45.548 | 0.185 | 0.027 | + | + | High |
| | | | | | 11T-SCM4 | 33.755 | 0.114 | 0.008 | + | + | Moderate |
| | | | | | 11T-SCM5 | 28.346 | 0.1556 | 0.011 | - | - | Moderate |
| | | | | | 11D-SCM1 | 13.74 | 0.236 | 0.019 | - | - | Moderate |
| | | | | | 11D-SCM2 | 12.225 | 0.194 | 0.01 | + | + | High |
| | | | | | 11D-SCM3 | 15.038 | 0.114 | 0.014 | - | - | Low |
| | 1 | 45966084 | MMACHC, c.609G>A | M | 11T-SCM1 | 14.497 | 0.137 | 0.019 | + | + | High |
| | | | | | 11T-SCM2 | 50.849 | 0.14 | 0.014 | + | + | Moderate |
| | | | | | 11T-SCM3 | 45.548 | 0.185 | 0.027 | + | + | High |
| | | | | | 11T-SCM4 | 33.755 | 0.114 | 0.008 | / | - | Undetermined |
| | | | | | 11T-SCM5 | 28.346 | 0.156 | 0.011 | - | - | Moderate |
| | | | | | 11D-SCM1 | 13.74 | 0.236 | 0.019 | - | - | High |
| | | | | | 11D-SCM2 | 12.225 | 0.194 | 0.01 | + | + | High |
| | | | | | 11D-SCM3 | 15.038 | 0.114 | 0.014 | - | - | Low |
| 12 | 6 | 49399569 | MUT, Exon13del | P | 12T-SCM1 | 50.849 | 0.131 | 0.013 | - | - | High |
| | | | | | 12D-SCM1 | 5.518 | 0.294 | 0.018 | - | - | High |
| | | | | | 12D-SCM2 | 19.799 | 0.109 | 0.012 | - | - | High |
| | | | | | 12D-SCM3 | 51.499 | 0.107 | 0.009 | / | + | Undetermined |
| | | | | | 12D-SCM4 | 12.009 | 0.147 | 0.023 | / | + | Rule Out |
| | 6 | 49419405 | MUT, c.1106G>A | M | 12T-SCM1 | 50.849 | 0.131 | 0.013 | - | - | Moderate |
| | | | | | 12D-SCM1 | 5.518 | 0.295 | 0.018 | - | - | Low |
| | | | | | 12D-SCM2 | 19.799 | 0.109 | 0.012 | + | + | High |
| | | | | | 12D-SCM3 | 51.499 | 0.107 | 0.009 | - | - | Low |
| | | | | | 12D-SCM4 | 12.009 | 0.147 | 0.023 | / | + | Rule Out |
| 13 | X | 153133901 - 153133911 | L1CAM, c.1549-1559del | M | 13T-SCM1 | 14.497 | 0.256 | 0.08 | / | + | Undetermined |
| | | | | | 13T-SCM2 | 40.896 | 0.318 | 0.059 | + | + | High |
| | | | | | 13T-SCM3 | 56.908 | 0.169 | 0.012 | + | + | Moderate |
| | | | | | 13T-SCM4 | 32.024 | 0.212 | 0.055 | - | - | High |
| | | | | | 13T-SCM5 | 59.937 | 0.142 | 0.015 | - | - | High |
| | | | | | 13T-SCM6 | 46.63 | 0.126 | 0.012 | - | - | High |
| 14 | X | 149832040 | MTM1,c.1602G>A | M | 14T-SCM1 | 49.01 | 0.133 | 0 | + | + | High |
| | | | | | 14T-SCM2 | 32.782 | 0.147 | 0 | - | - | High |
| | | | | | 14T-SCM3 | 28.238 | 0.146 | 0 | - | - | High |

| | | | | | | | | | | | |
|---|---|---|---|---|---|---|---|---|---|---|---|
| 15 | 13 | 32913558 | BRCA2, c.5067dupA | M | 15D-SCM1 | 81.388 | 0.138 | 0.013 | + | + | High |
| | | | | | 15T-SCM1 | 18.307 | 0.121 | 0.007 | / | + | Rule Out |
| | | | | | 15T-SCM2 | 83.721 | 0.101 | 0.005 | - | - | High |
| | | | | | 15T-SCM3 | 40.798 | 0.138 | 0.045 | - | - | Moderate |
| | | | | | 15T-SCM4 | 62.798 | 0.117 | 0.009 | - | - | Low |
| 16 | 17 | 48276948 | COL1A1, c.299-14_c.302del | M | 16D-SCM1 | 70.518 | 0.154 | 0.014 | + | + | High |
| | | | | | 16D-SCM2 | 56.779 | 0.147 | 0.013 | + | + | Low |
| | | | | | 16D-SCM3 | 54.091 | 0.123 | 0.012 | - | - | High |
| | | | | | 16T-SCM1 | 64.278 | 0.159 | 0.014 | - | - | High |
| | | | | | 16T-SCM2 | 46.808 | 0.148 | 0.015 | / | + | Undetermined |
| | | | | | 16T-SCM3 | 25.303 | 0.099 | 0.009 | / | - | Rule Out |
| | | | | | 16T-SCM4 | 34.354 | 0.124 | 0.010 | + | + | High |
| | | | | | 16T-SCM5 | 16.476 | 0.149 | 0.043 | / | - | Undetermined |
| | | | | | 16T-SCM6 | 62.933 | 0.158 | 0.010 | + | + | High |
| | | | | | 16T-SCM7 | 61.187 | 0.124 | 0.009 | - | - | Moderate |
| | | | | | 16T-SCM8 | 68.358 | 0.173 | 0.015 | - | - | Moderate |
| | | | | | 16T-SCM9 | 32.978 | 0.114 | 0.007 | + | + | High |
| | | | | | 16T-SCM10 | 31.024 | 0.144 | 0.011 | - | - | Low |
| | | | | | 16T-SCM11 | 32.051 | 0.149 | 0.013 | + | + | High |
| | | | | | 16T-SCM12 | 50.811 | 0.145 | 0.014 | + | + | Low |
| 17 | 17 | 29550493 | NF1, c.1756_1759del | M | 17D-SCM1 | 36.111 | 0.137 | 0.012 | + | + | Moderate |
| | | | | | 17D-SCM2 | 43.199 | 0.152 | 0.015 | - | - | Low |
| | | | | | 17D-SCM3 | 25.480 | 0.125 | 0.007 | + | + | Moderate |
| | | | | | 17T-SCM1 | 7.988 | 0.196 | 0.042 | / | - | Rule Out |
| | | | | | 17T-SCM2 | 37.265 | 0.151 | 0.010 | + | + | Low |
| | | | | | 17T-SCM3 | 44.191 | 0.151 | 0.011 | - | - | High |
| | | | | | 17T-SCM4 | 40.127 | 0.130 | 0.011 | - | - | High |
| | | | | | 17T-SCM5 | 29.347 | 0.125 | 0.012 | - | - | Low |
| | | | | | 17T-SCM6 | 32.204 | 0.117 | 0.009 | + | + | Low |
| | | | | | 17T-SCM7 | 23.038 | 0.134 | 0.010 | - | - | Low |
| | | | | | 17T-SCM8 | 23.096 | 0.138 | 0.011 | - | - | Low |

| | | | | | | | | | | | |
|---|---|---|---|---|---|---|---|---|---|---|---|
| 18 | 5 | 112173668 | APC, c.2378_2379del | P | 18D-SCM1 | 33.024 | 0. 114 | 0. 004 | + | + | High |
| | | | | | 18D-SCM2 | 47.163 | 0. 145 | 0. 007 | + | + | Low |
| | | | | | 18D-SCM3 | 53.084 | 0. 120 | 0. 008 | + | + | Low |
| | | | | | 18D-SCM4 | 41.621 | 0. 147 | 0. 007 | + | + | Low |
| | | | | | 18D-SCM5 | 52.507 | 0. 134 | 0. 004 | + | + | Low |
| | | | | | 18D-SCM6 | 74.761 | 0. 148 | 0. 006 | - | - | High |
| | | | | | 18D-SCM7 | 70.202 | 0. 141 | 0. 005 | - | - | High |
| | | | | | 18D-SCM8 | 52.116 | 0. 157 | 0. 007 | - | - | Moderate |
| | | | | | 18T-SCM1 | 16.284 | 0. 192 | 0. 011 | / | - | Rule Out |
| | | | | | 18T-SCM2 | 61.519 | 0. 146 | 0. 008 | + | + | High |
| | | | | | 18T-SCM3 | 44.406 | 0. 114 | 0. 005 | + | + | Low |
| | | | | | 18T-SCM4 | 79.065 | 0. 123 | 0. 007 | - | - | High |
| | | | | | 18T-SCM5 | 19.995 | 0. 125 | 0. 004 | + | + | Low |
| | | | | | 18T-SCM6 | 75.686 | 0. 122 | 0. 006 | + | + | Moderate |
| | | | | | 18T-SCM7 | 5.985 | 0. 354 | 0. 013 | / | - | Rule Out |
| | | | | | 18T-SCM8 | 66.202 | 0. 139 | 0. 006 | + | + | High |
| 19 | 16 | 2140782 | PKD1, c.12031C>T | P | 19D-SCM1 | 30.246 | 0. 124 | 0. 007 | + | + | Moderate |
| | | | | | 19D-SCM2 | 24.804 | 0. 092 | 0. 004 | - | - | Low |
| | | | | | 19T-SCM1 | 13.931 | 0. 125 | 0. 008 | / | + | Rule Out |
| | | | | | 19T-SCM2 | 29.362 | 0. 107 | 0. 006 | - | - | Low |
| | | | | | 19T-SCM3 | 48.520 | 0. 126 | 0. 008 | + | + | Moderate |
| 20 | 7 | 107323898 | SLC26A4, c.919-2A>G | M | 20T-SCM1 | 48.944 | 0. 113 | 0. 034 | - | - | Low |
| | | | | | 20T-SCM2 | 5.950 | 0. 463 | 0. 521 | / | - | Rule Out |
| | | 107323898 | SLC26A4, c.919-2A>G | P | 20T-SCM1 | 48.944 | 0. 113 | 0. 034 | - | - | High |
| | | | | | 20T-SCM2 | 5.950 | 0. 463 | 0. 521 | / | - | Rule Out |
| 21 | 12 | 103246707 | PAH, c.728G>A | M | 21T-SCM1 | 35.595 | 0. 120 | 0. 007 | + | + | High |
| | | | | | 21T-SCM2 | 33.543 | 0. 122 | 0. 036 | + | + | High |
| | | | | | 21T-SCM3 | 51.858 | 0. 118 | 0. 006 | + | + | Moderate |
| | | | | | 21T-SCM4 | 51.597 | 0. 102 | 0. 006 | - | - | Low |
| | | 103246707 | PAH, c.728G>A | P | 21T-SCM1 | 35.595 | 0. 120 | 0. 007 | + | + | Low |
| | | | | | 21T-SCM2 | 33.543 | 0. 122 | 0. 036 | + | + | Moderate |
| | | | | | 21T-SCM3 | 51.858 | 0. 118 | 0. 006 | + | + | High |
| | | | | | 21T-SCM4 | 51.597 | 0. 102 | 0. 006 | / | + | Undetermined |

| | | | | | | | | | | | |
|---|---|---|---|---|---|---|---|---|---|---|---|
| 22 | 17 | 41052814 | G6PC1, exon1-2del | P | 22T-SCM1 | 22.291 | 0.107 | 0.012 | - | - | Low |
| | | | | | 22T-SCM2 | 53.191 | 0.128 | 0.028 | - | - | High |
| | | | | | 22T-SCM3 | 3.419 | 0.629 | 0.180 | / | + | Rule Out |
| | | | | | 22T-SCM4 | 53.837 | 0.128 | 0.026 | - | - | Moderate |
| | | 41063017 | G6PC1, c.648G>T | M | 22T-SCM1 | 22.291 | 0.107 | 0.012 | / | - | Undetermined |
| | | | | | 22T-SCM2 | 53.191 | 0.128 | 0.028 | - | - | High |
| | | | | | 22T-SCM3 | 3.419 | 0.629 | 0.180 | / | + | Rule Out |
| | | | | | 22T-SCM4 | 53.837 | 0.128 | 0.026 | + | + | Low |
| 23 | 12 | 103237426 | PAH, c.1197A>T | M | 23D-SCM1 | 35.755 | 0.123 | 0.006 | - | - | High |
| | | | | | 23D-SCM2 | 42.421 | 0.151 | 0.006 | + | + | Moderate |
| | | | | | 23D-SCM3 | 24.538 | 0.141 | 0.007 | + | + | Low |
| | | | | | 23D-SCM4 | 35.353 | 0.141 | 0.009 | - | - | Moderate |
| | | | | | 23D-SCM5 | 70.724 | 0.143 | 0.008 | - | - | High |
| | | | | | 23T-SCM1 | 37.731 | 0.168 | 0.008 | + | + | Low |
| | | | | | 23T-SCM2 | 10.679 | 0.192 | 0.012 | + | + | Moderate |
| | | 103249088 | PAH, c.532G>A | P | 23D-SCM1 | 35.755 | 0.123 | 0.006 | + | + | Low |
| | | | | | 23D-SCM2 | 42.421 | 0.151 | 0.006 | + | + | High |
| | | | | | 23D-SCM3 | 24.538 | 0.141 | 0.007 | + | + | High |
| | | | | | 23D-SCM4 | 35.353 | 0.141 | 0.009 | + | + | Low |
| | | | | | 23D-SCM5 | 70.724 | 0.143 | 0.008 | + | + | High |
| | | | | | 23T-SCM1 | 37.731 | 0.168 | 0.008 | + | + | High |
| | | | | | 23T-SCM2 | 10.679 | 0.192 | 0.012 | + | + | High |

| | | | | | | | | | | | |
|---|---|---|---|---|---|---|---|---|---|---|---|
| 24 | 1 | 216498798 | USH2A, c.991dupA | P | 24D-SCM1 | 15.097 | 0. 129 | 0. 016 | + | + | High |
| | | | | | 24D-SCM2 | 14.795 | 0. 116 | 0. 019 | - | - | High |
| | | | | | 24D-SCM3 | 4.860 | 0. 180 | 0. 012 | + | + | Low |
| | | | | | 24D-SCM4 | 7.409 | 0. 122 | 0. 011 | + | + | Low |
| | | | | | 24D-SCM5 | 14.696 | 0. 144 | 0. 017 | + | + | High |
| | | | | | 24T-SCM1 | 35.125 | 0. 105 | 0. 016 | - | - | Moderate |
| | | | | | 24T-SCM2 | 5.239 | 0. 097 | 0. 011 | + | + | Low |
| | | | | | 24T-SCM3 | 12.876 | 0. 087 | 0. 009 | + | + | Moderate |
| | | 216595579 | USH2A, c.99_100insT | M | 24D-SCM1 | 15.097 | 0. 129 | 0. 016 | - | - | Low |
| | | | | | 24D-SCM2 | 14.795 | 0. 116 | 0. 019 | + | + | High |
| | | | | | 24D-SCM3 | 4.860 | 0. 180 | 0. 012 | / | + | Undetermined |
| | | | | | 24D-SCM4 | 7.409 | 0. 122 | 0. 011 | - | - | High |
| | | | | | 24D-SCM5 | 14.696 | 0. 144 | 0. 017 | / | + | Undetermined |
| | | | | | 24T-SCM1 | 35.125 | 0. 105 | 0. 016 | - | - | Moderate |
| | | | | | 24T-SCM2 | 5.239 | 0. 097 | 0. 011 | + | + | Low |
| | | | | | 24T-SCM3 | 12.876 | 0. 087 | 0. 009 | + | + | Moderate |
| 25 | 2 | 219674454 | CYP27A1, c.410G>A | M | 25T-SCM1 | 66.732 | 0. 122 | 0. 012 | + | + | Low |
| | | | | | 25T-SCM2 | 17.009 | 0. 102 | 0. 011 | / | - | Undetermined |
| | | | | | 25T-SCM3 | 15.143 | 0. 101 | 0. 015 | / | + | Undetermined |
| | | | | | 25T-SCM4 | 28.686 | 0. 095 | 0. 011 | - | - | Low |
| | | | | | 25T-SCM5 | 36.507 | 0. 128 | 0. 008 | - | - | Low |
| | | | | | 25T-SCM6 | 13.741 | 0. 307 | 0. 016 | - | - | High |
| | | | | | 25T-SCM7 | 21.498 | 0. 290 | 0. 014 | / | + | Rule Out |
| | | | | | 25T-SCM8 | 13.783 | 0. 249 | 0. 015 | - | - | Moderate |
| | | | | | 25T-SCM9 | 19.802 | 0. 172 | 0. 015 | / | + | Rule Out |
| | | | | | 25T-SCM10 | 20.348 | 0. 178 | 0. 013 | / | + | Undetermined |
| | | 219674454 | CYP27A1, c.410G>A | P | 25T-SCM1 | 66.732 | 0. 122 | 0. 012 | + | + | Moderate |
| | | | | | 25T-SCM2 | 17.009 | 0. 102 | 0. 011 | - | - | Low |
| | | | | | 25T-SCM3 | 15.143 | 0. 101 | 0. 015 | + | + | Low |
| | | | | | 25T-SCM4 | 28.686 | 0. 095 | 0. 011 | - | - | High |
| | | | | | 25T-SCM5 | 36.507 | 0. 128 | 0. 008 | - | - | High |
| | | | | | 25T-SCM6 | 13.741 | 0. 307 | 0. 016 | - | - | Moderate |
| | | | | | 25T-SCM7 | 21.498 | 0. 290 | 0. 014 | / | + | Rule Out |
| | | | | | 25T-SCM8 | 13.783 | 0. 249 | 0. 015 | + | + | High |
| | | | | | 25T-SCM9 | 19.802 | 0. 172 | 0. 015 | / | - | Rule Out |
| | | | | | 25T-SCM10 | 20.348 | 0. 178 | 0. 013 | - | - | Low |

| | | | | | | | | | | | |
|---|---|---|---|---|---|---|---|---|---|---|---|
| 26 | 2 | 169830269 | CB11, c.1389_1390insT | P | 26T-SCM1 | 78.125 | 0.120 | 0.015 | - | - | Moderate |
| | | | | | 26T-SCM2 | 59.125 | 0.114 | 0.012 | + | + | High |
| | | | | | 26T-SCM3 | 58.870 | 0.126 | 0.016 | - | - | High |
| | | | | | 26T-SCM4 | 43.381 | 0.115 | 0.011 | / | - | Undetermined |
| | | 169842713 | ABCB11, c.990G>C | M | 26T-SCM1 | 78.125 | 0.120 | 0.015 | + | + | High |
| | | | | | 26T-SCM2 | 59.125 | 0.114 | 0.012 | + | + | High |
| | | | | | 26T-SCM3 | 58.870 | 0.126 | 0.016 | + | + | High |
| | | | | | 26T-SCM4 | 43.381 | 0.115 | 0.011 | + | + | Low |
| 27 | 13 | 20763486 | GJB2, c.235delC | M | 27T-SCM1 | 20.881 | 0.131 | 0.007 | / | - | Low |
| | | | | | 27T-SCM2 | 54.600 | 0.131 | 0.006 | + | + | Low |
| | | | | | 27T-SCM3 | 41.165 | 0.108 | 0.004 | + | + | High |
| | | | | | 27T-SCM4 | 30.098 | 0.137 | 0.006 | / | + | Low |
| | | | | | 27T-SCM5 | 31.705 | 0.108 | 0.006 | / | - | Low |
| | | 20763486 | GJB2, c.235delC | P | 27T-SCM1 | 20.881 | 0.131 | 0.007 | + | + | Low |
| | | | | | 27T-SCM2 | 54.600 | 0.131 | 0.006 | + | + | High |
| | | | | | 27T-SCM3 | 41.165 | 0.108 | 0.004 | / | - | Undetermined |
| | | | | | 27T-SCM4 | 30.098 | 0.137 | 0.006 | - | - | Moderate |
| | | | | | 27T-SCM5 | 31.705 | 0.108 | 0.006 | + | + | High |
| 28 | 13 | 20763421 | GJB2, c.299delAT | P | 28T-SCM1 | 48.152 | 0.115 | 0.009 | - | - | Low |
| | | | | | 28T-SCM2 | 25.847 | 0.114 | 0.008 | + | + | Moderate |
| | | 20763486 | GJB2, c.235delC | M | 28T-SCM1 | 48.152 | 0.115 | 0.009 | + | + | High |
| | | | | | 28T-SCM2 | 25.847 | 0.114 | 0.008 | + | + | Low |
| 29 | 13 | 20763612 | GJB2, c.109G>A | M | 29T-SCM1 | 38.140 | 0.137 | 0.007 | - | - | Moderate |
| | | | | | 29T-SCM2 | 24.561 | 0.143 | 0.004 | + | + | Moderate |
| | | | | | 29T-SCM3 | 17.547 | 0.179 | 0.007 | / | + | Rule Out |
| | | | | | 29T-SCM4 | 24.350 | 0.170 | 0.006 | + | + | Low |
| | | 20763612 | GJB2, c.109G>A | P | 29T-SCM1 | 38.140 | 0.137 | 0.007 | + | + | Moderate |
| | | | | | 29T-SCM2 | 24.561 | 0.143 | 0.004 | + | + | High |
| | | | | | 29T-SCM3 | 17.547 | 0.179 | 0.007 | / | - | Rule Out |
| | | | | | 29T-SCM4 | 24.350 | 0.170 | 0.006 | - | - | Low |

#### Table S6. Source data for statistics in Fig. 3D and 3E.

| Sample | Coverage before imputation | Coverage after imputatioin |
|---|---|---|
| 1T-SCM1 | 0.0827 | 0.9607 |
| 1T-SCM2 | 0.1064 | 0.9657 |
| 1T-SCM3 | 0.0651 | 0.9472 |
| 1T-SCM4 | 0.0636 | 0.935 |
| 1T-SCM5 | 0.0551 | 0.9401 |
| 1T-SCM6 | 0.0166 | 0.8303 |
| 2T-SCM1 | 0.0136 | 0.6837 |
| 2T-SCM2 | 0.0103 | 0.8495 |
| 2T-SCM3 | 0.0194 | 0.7962 |
| 2T-SCM4 | 0.0094 | 0.7795 |
| 2T-SCM5 | 0.0063 | 0.7523 |
| 2T-SCM6 | 0.0115 | 0.8768 |
| 2T-SCM7 | 0.0123 | 0.875 |
| 2T-SCM8 | 0.0065 | 0.8269 |
| 3T-SCM1 | 0.1419 | 0.9642 |
| 3T-SCM2 | 0.0827 | 0.9486 |
| 3T-SCM3 | 0.1173 | 0.9759 |
| 3T-SCM4 | 0.138 | 0.9661 |
| 3T-SCM5 | 0.1842 | 0.9824 |
| 3D-SCM2 | 0.1141 | 0.8692 |
| 3D-SCM3 | 0.1116 | 0.9587 |
| 4T-SCM1 | 0.0047 | 0.5363 |
| 4T-SCM2 | 0.012 | 0.9406 |
| 4T-SCM3 | 0.0064 | 0.8214 |
| 4T-SCM4 | 0.0008 | 0.4933 |
| 4T-SCM5 | 0.0022 | 0.7814 |
| 4T-SCM6 | 0.0024 | 0.825 |
| 4T-SCM7 | 0.0068 | 0.8301 |
| 4D-SCM1 | 0.0041 | 0.8526 |
| 4D-SCM2 | 0.0017 | 0.6468 |
| 4D-SCM3 | 0.0059 | 0.8285 |
| 4D-SCM4 | 0.0075 | 0.882 |
| 5T-SCM1 | 0.0669 | 0.9579 |
| 5T-SCM2 | 0.1102 | 0.9565 |
| 5D-SCM2 | 0.0631 | 0.9196 |
| 6T-SCM1 | 0.0753 | 0.958 |
| 6T-SCM2 | 0.1486 | 0.9688 |
| 6D-SCM2 | 0.1217 | 0.9911 |
| 7T-SCM1 | 0.0996 | 0.9582 |
| 7T-SCM2 | 0.1243 | 0.9763 |
| 7D-SCM1 | 0.1245 | 0.9551 |
| 8T-SCM1 | 0.1485 | 0.9851 |
| 8T-SCM2 | 0.0173 | 0.9622 |
| 8T-SCM3 | 0.012 | 0.939 |
| 8T-SCM4 | 0.0211 | 0.9215 |
| 8T-SCM5 | 0.1653 | 0.9276 |
| 9T-SCM1 | 0.0014 | 0.6855 |
| 9T-SCM2 | 0.0062 | 0.8181 |
| 9T-SCM3 | 0.0067 | 0.8046 |
| 9T-SCM4 | 0.0024 | 0.6806 |
| 9T-SCM5 | 0.2777 | 0.9814 |
| 9T-SCM6 | 0.0644 | 0.9729 |
| 9T-SCM7 | 0.0233 | 0.8909 |

| | | |
|---|---|---|
| 9T-SCM8 | 0.0123 | 0.9104 |
| 9T-SCM9 | 0.03 | 0.9562 |
| 9D-SCM1 | 0.1292 | 0.9701 |
| 9D-SCM2 | 0.0035 | 0.8802 |
| 9D-SCM3 | 0.1202 | 0.9437 |
| 9D-SCM4 | 0.1292 | 0.9606 |
| 10T-SCM1 | 0.1312 | 0.9671 |
| 10T-SCM2 | 0.0712 | 0.8725 |
| 10T-SCM3 | 0.149 | 0.9762 |
| 10D-SCM1 | 0.1486 | 0.9665 |
| 10D-SCM2 | 0.2006 | 0.9738 |
| 11T-SCM1 | 0.0457 | 0.9007 |
| 11T-SCM2 | 0.1084 | 0.9881 |
| 11T-SCM3 | 0.0687 | 0.9818 |
| 11T-SCM4 | 0.1121 | 0.9868 |
| 11T-SCM5 | 0.0605 | 0.9296 |
| 11D-SCM1 | 0.0404 | 0.9116 |
| 11D-SCM2 | 0.0399 | 0.9355 |
| 11D-SCM3 | 0.0521 | 0.8949 |
| 12T-SCM1 | 0.1439 | 0.9829 |
| 12D-SCM1 | 0.0147 | 0.8631 |
| 12D-SCM2 | 0.0738 | 0.9678 |
| 12D-SCM3 | 0.1452 | 0.9847 |
| 12D-SCM4 | 0.0395 | 0.7038 |
| 13T-SCM1 | 0.0191 | 0.9119 |
| 13T-SCM2 | 0.0123 | 0.9218 |
| 13T-SCM3 | 0.0989 | 0.9726 |
| 13T-SCM4 | 0.0195 | 0.8996 |
| 13T-SCM5 | 0.1375 | 0.9811 |
| 13T-SCM6 | 0.134 | 0.9747 |
| 14T-SCM1 | 0.1181 | 0.974 |
| 14T-SCM2 | 0.0811 | 0.9659 |
| 14T-SCM3 | 0.0891 | 0.9649 |

| Sample | High Maternal Contamined | Concordance of Genotype | Concordance of Maternal Haplotype | Concordance of Paternal Haplotype |
|---|---|---|---|---|
| 1T-SCM1 | no | 0.9484 | 0.9996 | 0.9984 |
| 1T-SCM2 | no | 0.9471 | 0.9993 | 0.9982 |
| 1T-SCM3 | no | 0.9469 | 0.9999 | 0.999 |
| 1T-SCM4 | no | 0.9447 | 0.9998 | 0.9983 |
| 1T-SCM6 | no | 0.951 | 0.9961 | 0.9932 |
| 2T-SCM1 | no | 0.9677 | 0.9296 | 0.9986 |
| 2T-SCM2 | no | 0.9728 | 0.9857 | 0.9991 |
| 2T-SCM4 | no | 0.9733 | 0.9765 | 0.9964 |
| 2T-SCM5 | no | 0.9733 | 0.9782 | 0.9975 |
| 2T-SCM6 | no | 0.9734 | 0.9966 | 0.9975 |
| 2T-SCM7 | no | 0.9718 | 0.9973 | 0.9983 |
| 2T-SCM8 | no | 0.9739 | 0.9898 | 0.9955 |
| 3T-SCM1 | no | 0.9814 | 0.972 | 0.9993 |
| 3T-SCM2 | no | 0.8992 | 0.7593 | 1 |
| 3T-SCM3 | no | 0.9834 | 0.986 | 1 |
| 3T-SCM4 | no | 0.9785 | 0.9913 | 1 |
| 3D-SCM3 | no | 0.977 | 0.9782 | 0.9999 |
| 4D-SCM3 | no | 0.9882 | 0.9918 | 0.984 |
| 7T-SCM1 | no | 0.9436 | 0.9888 | 0.9928 |
| 7D-SCM1 | no | 0.9107 | 0.9867 | 0.9947 |
| 8T-SCM4 | no | 0.9939 | 0.9672 | 1 |
| 8T-SCM5 | no | 0.9957 | 0.9728 | 0.9976 |
| 9T-SCM5 | no | 0.9994 | 1 | 0.9999 |
| 9T-SCM7 | no | 0.9849 | 0.958 | 0.9953 |
| 9T-SCM9 | no | 0.9993 | 1 | 0.9999 |
| 9D-SCM1 | no | 0.8986 | 0.9985 | 0.9932 |
| 10T-SCM1 | no | 0.9926 | 0.9939 | 0.9977 |
| 10T-SCM2 | yes | 0.7921 | 0.3874 | 0.9923 |
| 10D-SCM1 | no | 0.9747 | 0.941 | 0.9999 |
| 10D-SCM2 | no | 0.9903 | 0.9801 | 0.9997 |
| 11T-SCM1 | yes | 0.8673 | 0.3783 | 1 |
| 11T-SCM3 | no | 0.9994 | 0.9862 | 1 |
| 11T-SCM5 | no | 0.9723 | 0.7689 | 0.9999 |
| 11D-SCM1 | no | 0.9407 | 0.8221 | 0.8646 |
| 12D-SCM1 | no | 0.9884 | 0.9828 | 0.995 |
| 12D-SCM4 | yes | 0.8249 | 0.2842 | 0.6831 |
| 13T-SCM1 | no | 0.965 | 0.9771 | 0.9928 |
| 13T-SCM2 | no | 0.971 | 0.9988 | 0.9982 |
| 13T-SCM3 | no | 0.9658 | 0.9967 | 0.9993 |
| 13T-SCM4 | no | 0.9681 | 0.9905 | 0.9967 |
| 13T-SCM6 | no | 0.9713 | 0.9998 | 0.9996 |
| 14T-SCM1 | no | 0.9898 | 0.9821 | 1 |
| 14T-SCM2 | no | 0.9966 | 0.9983 | 0.9981 |

#### Table S7. Source data for statistics in Fig. 4.

| Sample | Directly detected SNPs | Pedigree-based imputed SNPs | Population-based imputed SNPs |
|---|---|---|---|
| 1T-SCM4 | 35 | 67 | 12 |
| 2T-SCM2 | 3 | 87 | 24 |
| 2T-SCM4 | 9 | 76 | 29 |
| 2T-SCM5 | 6 | 77 | 31 |
| 2T-SCM6 | 5 | 85 | 24 |
| 2T-SCM8 | 11 | 77 | 26 |
| 3T-SCM1 | 44 | 47 | 23 |
| 3T-SCM3 | 60 | 30 | 24 |
| 3T-SCM4 | 36 | 51 | 27 |
| 3D-SCM3 | 25 | 64 | 25 |
| 7T-SCM1 | 26 | 78 | 10 |
| 7D-SCM1 | 38 | 69 | 7 |
| 8T-SCM4 | 29 | 23 | 62 |
| 9T-SCM5 | 72 | 20 | 22 |
| 9T-SCM7 | 32 | 55 | 27 |
| 9T-SCM9 | 38 | 51 | 25 |
| 9D-SCM1 | 34 | 58 | 22 |
| 10D-SCM1 | 57 | 47 | 10 |
| 10D-SCM2 | 53 | 52 | 9 |
| 11T-SCM3 | 38 | 57 | 19 |
| 11T-SCM5 | 22 | 70 | 22 |
| 13T-SCM2 | 27 | 64 | 23 |
| 13T-SCM4 | 28 | 72 | 14 |
| 13T-SCM6 | 39 | 62 | 13 |
| 14T-SCM1 | 50 | 58 | 6 |

| Sample | 0/0 identical sites | 1/1 identical sites | 0/1 identical sites |
|---|---|---|---|
| 1T-SCM4 | 59 | 21 | 31 |
| 2T-SCM2 | 64 | 25 | 20 |
| 2T-SCM4 | 60 | 25 | 18 |
| 2T-SCM5 | 58 | 23 | 25 |
| 2T-SCM6 | 65 | 22 | 21 |
| 2T-SCM8 | 60 | 20 | 26 |
| 3T-SCM1 | 55 | 30 | 28 |
| 3T-SCM3 | 56 | 28 | 30 |
| 3T-SCM4 | 62 | 31 | 17 |
| 3D-SCM3 | 55 | 31 | 27 |
| 7T-SCM1 | 64 | 26 | 19 |
| 7D-SCM1 | 62 | 29 | 22 |
| 8T-SCM4 | 53 | 28 | 28 |
| 9T-SCM5 | 47 | 26 | 12 |
| 9T-SCM7 | 49 | 25 | 16 |
| 9T-SCM9 | 49 | 24 | 16 |
| 9D-SCM1 | 56 | 29 | 27 |
| 10D-SCM1 | 56 | 30 | 26 |
| 10D-SCM2 | 58 | 31 | 25 |
| 11T-SCM3 | 56 | 26 | 29 |
| 11T-SCM5 | 58 | 27 | 21 |
| 13T-SCM2 | 56 | 26 | 32 |
| 13T-SCM4 | 51 | 30 | 32 |
| 13T-SCM6 | 63 | 26 | 24 |
| 14T-SCM1 | 49 | 29 | 34 |

| Sample | PRS_CM | Percentile_CM | PRS_gold_standard | Percentile_gold_standard | Group |
|---|---|---|---|---|---|
| 1T-SCM4 | 16.50539 | 42.42062501 | 16.77061565 | 58.01326022 | TE |
| 2T-SCM2 | 15.67084 | 7.395501026 | 15.79465677 | 10.41406452 | TE |
| 2T-SCM4 | 15.98312 | 16.54988554 | 15.97123664 | 16.09593389 | TE |
| 2T-SCM5 | 15.67918 | 7.581013068 | 15.65245776 | 7.007686669 | TE |
| 2T-SCM6 | 15.46502 | 3.898209993 | 15.59225434 | 5.865743338 | TE |
| 2T-SCM8 | 15.69986 | 8.047250286 | 16.00414194 | 17.35397581 | TE |
| 3T-SCM1 | 16.47932 | 40.93509536 | 16.35499492 | 33.92699917 | TE |
| 3T-SCM3 | 16.9034 | 65.46056538 | 16.90340082 | 65.46056538 | TE |
| 3T-SCM4 | 17.03899 | 72.5734984 | 16.89950081 | 65.25478216 | TE |
| 3D-SCM3 | 16.99581 | 70.42598151 | 16.95316975 | 68.19328471 | discard |
| 7T-SCM1 | 16.72671 | 55.47895278 | 16.72330427 | 55.26723153 | TE |
| 7D-SCM1 | 16.32404 | 32.26394309 | 16.14771454 | 23.46768282 | discard |
| 8T-SCM4 | 16.52919 | 43.83366982 | 16.72595926 | 55.42858196 | TE |
| 9T-SCM5 | 17.00268 | 70.76260601 | 16.95657558 | 68.37060637 | TE |
| 9T-SCM7 | 16.6431 | 50.57824063 | 16.95657558 | 68.37060637 | TE |
| 9T-SCM9 | 16.29637 | 30.80912736 | 16.95657558 | 68.37060637 | TE |
| 9D-SCM1 | 15.80035 | 10.57971489 | 16.11208751 | 21.8547928 | discard |
| 10D-SCM1 | 16.17982 | 24.98024072 | 16.37504027 | 35.02369067 | discard |
| 10D-SCM2 | 16.8807 | 64.22975646 | 16.88069923 | 64.22975646 | discard |
| 11T-SCM3 | 16.29103 | 30.52307844 | 16.54199239 | 44.59476062 | TE |
| 11T-SCM5 | 17.07885 | 74.49250376 | 16.37373967 | 34.9559153 | TE |
| 13T-SCM2 | 17.14947 | 77.69698881 | 17.14947373 | 77.69698881 | TE |
| 13T-SCM4 | 16.40653 | 36.78871694 | 16.37796352 | 35.18115067 | TE |
| 13T-SCM6 | 16.8009 | 59.75146301 | 16.91193092 | 65.91513131 | TE |
| 14T-SCM1 | 15.01846 | 0.717886555 | 15.04517752 | 0.802452199 | TE |
| 1T-SCM3 | 16.50138 | 42.19948483 | 16.82292724 | 61.01278108 | AmnioticFluid |
| 3T-SCM1 | 16.47932 | 40.93509536 | 16.31460736 | 31.76924431 | AmnioticFluid |
